# Self-reported health history from 70,724 individuals reveals novel HLA associations with allergy and other frequently underreported conditions

**DOI:** 10.64898/2026.02.18.26346586

**Authors:** Juliano André Boquett, Sean Yu-Tseng Lin, John S. House, Kwangmi Ahn, Rayo Suseno, Abena BakenRa, Karoline Guthrie, Michael Wright, Alison A. Motsinger-Reif, Martin Maiers, Jill A. Hollenbach

## Abstract

**Background:** Variation in the *HLA* loci, located on human chromosome 6p, has been associated with hundreds of diseases and conditions. However, high levels of polymorphism that characterize the *HLA* system, coupled with generally modest effect sizes for most phenotypes, necessitate relatively large sample sizes to power association studies; meanwhile, high resolution HLA genotyping remains relatively resource intensive. These constraints limit identification of novel associations. While phenome-wide association studies (PheWAS) in the context of large registries with available electronic health records (EHR) have revealed new insights into the role of *HLA* in disease, many common health conditions are poorly represented in EHR due to the temporal nature of their occurrence or general underreporting. Further, these studies have generally been conducted with *HLA* genotyping data imputed from microarrays, rather than direct measurement of high-resolution genotypes.

**Objective:** To overcome these limitations and reveal novel *HLA* associations we undertook a PheWAS in many previously understudied health conditions.

**Methods:** We queried over 300 hundred conditions, diseases and traits from 70,724 subjects registered with NMDP with available high-resolution *HLA* genotyping (*HLA-A*, *HLA-B*, *HLA-C*, *HLA-DRB1*, and *HLA-DQB1*). After stratifying according to ancestry, we performed a logistic regression analysis adjusting for sex and age for HLA-phenotype association.

**Results:** We identified 48 significant *HLA* associations across ancestry groups, confirming several known associations and uncovered fifteen novel associations. Most novel associations pertained to common infectious or allergic phenotypes that often go under-reported in the EHR. Of particular translational importance, we identified a previously undetected yet very strong association between HLA-DRB1*04:01 and sensitivity to cefaclor, a specific class of cephalosporin (OR = 3.74, p-value 5.10E-28). Molecular docking simulations predict cefaclor binding in the P4 pocket of HLA-DRB1*04:01, with substantially greater affinity than non-associated antibiotics, including other cephalosporins. This pharmacogenomic signal highlights an opportunity for risk stratification and targeted prevention of adverse drug reactions. Other novel associations found, such as susceptibility to genital warts (HPV) and allergic rhinitis, reveals new insights into the role of specific *HLA* alleles in immune-mediated disease. The vast majority of these novel associations were replicated in the independent All of Us cohort, confirming the validity of this approach.

**Conclusion:** Collectively, our findings demonstrate the value of integrating population-scale, high-resolution *HLA* genotypes with phenotyping beyond the EHR to reveal immunogenetic influences on common health outcomes. They also point to immediate translational avenues – particularly for drug hypersensitivity – while motivating future functional studies and prospective clinical validation to refine mechanistic understanding and clinical utility.

## Introduction

*HLA* (Human Leukocyte Antigen, chr 6p21) are the most polymorphic genes of the human genome and the most medically significant. Variation at *HLA* has long been associated with human health, including in autoimmune and infectious diseases, in drug hypersensitivities and in a range of immune-mediated phenotypes(1–3). The *HLA* genes encode cell-surface molecules whose role is to present antigen, both foreign and self-derived, to T cells(4–6). The high levels of sequence variability in the *HLA* genes results in exquisite specificity with regard to antigen binding and interaction with the T cell receptor(7–10).

Phenome-wide association studies (PheWAS), a genotype-to-phenotype strategy, have been extremely successful in uncovering novel genetic associations. Studies exploiting existing genetic data in the context of large public biorepositories, and linked to electronic health records (EHR), have allowed orders of magnitude increases in statistical power to discover unexplored and cross-phenotype associations(11–16). Most PheWAS to date have examined genetic variants against phenotypes recorded in EHR(17); these have revealed known and novel genetic associations across the genome, including at *HLA*(11,14,16). Nevertheless, many phenotypes are under-reported in EHR, such as early childhood infections, sexually transmitted diseases, and behavioral health information(18–22). Despite decades of research inspecting the association between *HLA* variation and disease, many common phenotypes have been largely ignored, limiting a comprehensive understanding of *HLA*’s influence on human health.

To close this knowledge gap, we undertook an *HLA* PheWAS aimed at uncovering novel HLA associations with health outcomes that may be underrepresented in the EHR. Leveraging existing high-resolution *HLA* genotyping and collecting, through self-report, wide-ranging health data specifically targeting common medical conditions not consistently noted in medical records, we examined more than 300 hundred phenotypes from more than 70,000 individuals registered as potential bone marrow donors with the NMDP (formerly known as the National Marrow Donor Program) with available high-resolution *HLA* genotyping data. To confirm novel associations, we utilized genetic data from the All of Us (AoU) research program, encompassing 253,128 individuals with both EHR and genetic data. Our results support numerous previously reported *HLA*-phenotype associations while revealing 15 novel *HLA* associations, primarily related to infection and allergy, including a strong and significant risk for antibiotic hypersensitivity.

## Methods

### NMDP (discovery) cohort

In January 2023, all registered potential volunteer bone marrow donors in the NMDP database with available email addresses were invited to participate in the study via email outreach. The email provided a custom link directing them to a consent page in our health history survey. Individuals who lacked high resolution *HLA* data and those who completed the survey multiple times were excluded from the study.

The final discovery cohort consisted of anonymized data from 70,724 participants with the following self-reported ancestry: 61,825 European descent (EUR), 5,236 Hispanic or Latino (HIS), 2,536 Asian or Pacific Islander (API), 1,127 African American or Black (AFA). A total of 7,026 individuals comprising the self-reported ancestries as Native American (NAM), multiracial (MLT), unknown ancestry (UNK), identified as other (OTH) or declined to specify (DEC) were not included in the analysis due to small sample size and/or due to prevent confounding. Detailed demographic characteristics including ancestries, sex and age are shown in Table 1.

**Table 1.** Demographic characteristics for the discovery (NMDP) cohort.

| **Ancestry** | **N** | **Female %** | **Mean age (SD)** |
| --- | --- | --- | --- |
| African American (AFA) | 1,127 | 83.5 | 41.4 (10.1) |
| Asian or Pacific Islander (API) | 2,536 | 68.8 | 39.2 (9.6) |
| European descent (EUR) | 61,825 | 79.9 | 39.6 (10.1) |
| Hispanic or Latino (HIS) | 5,236 | 79.3 | 37.9 (9.8) |

High-resolution *HLA* genotyping from the registry database for classical class I (*HLA-A*, *HLA-B*, *HLA-C*) and class II (*HLA-DRB1* and *HLA-DQB1*) loci was linked to survey responses. Genotyping was previously performed upon donor recruitment through next-generation sequencing (NGS), covering the antigen recognition domain (ARD) exons. The PacBio sequencing platform was used to perform full gene-phased sequencing. Any genotyping ambiguities are addressed through a bioinformatic method aimed at resolving these genotypes(23). The data were deidentified and use for this study was approved by the UCSF IRB (Institutional Review Board).

### Health History Survey

Subject recruitment, consent, and survey administration were conducted electronically via email and a web interface, ensuring efficient and comprehensive data collection. Every participant in the study provided written informed consent. Participants were asked to complete a detailed health history survey, taking approximately 10 to 15 minutes. The health history survey was developed to capture data for a wide range of common conditions that may be absent or under-reported in the EHR. To conform to previously validated surveys, we followed the naming conventions for lay terms and survey methods and resources developed and validated under the Clinical Genome Resource (ClinGen) initiative(24). The survey included a total of 357 phenotypes including autoimmune diseases, allergies, heart conditions, mental health and behavioral health symptoms, neurological disorders, cancer, skin conditions, digestive disorders, infectious diseases, and pregnancy conditions.

Each phenotype was coded as a binary variable in our dataset. We also queried frequency of certain infectious diseases (e.g., strep throat, flu, bronchitis, pneumonia, and ear infections during childhood) was categorized as: ‘rarely’ for occurrences every five years or more, ‘occasionally’ for once every 2-5 years, ‘frequently’ for an annual occurrence, and ‘very frequently’ for more than once per year; these were coded as ordinal variables. In addition, the survey featured free-text fields designed to capture detailed information for each category, including food and drug allergies. We examined these open-ended responses for keywords indicating specific allergies, such as “penicillin” or “PNC.” Once identified, these specifics were converted into binary variables.

### All of Us (replication) cohort

The AoU research program database was used as a replication cohort for novel associations found in the discovery cohort. For analyses, the AoU cohort restricted to unrelated individuals with a defined sex at birth, short read WGS (srWGS) and EHR data who had consented for research(25). This analysis included participants of African (AFR; n = 49,410), admixed American (AMR; n = 46,916) and European (EUR; n = 156,802) ancestries. *HLA* genotyping was performed using srWGS data and Kourami v0.9.6(26). For mapping ICD-9 and ICE-10 codes to phecodes, we employed PhecodeX 1.0 from the PheTK package. In brief, individuals with zero EHR records were coded as control, those with two or more records at distinct time points were coded as cases, and individuals with only a single record were not considered. Phecodes with fewer than 50 cases were not considered(27).

### HLA association analysis

We examined the association of five *HLA* loci (*HLA-A*, *HLA-B*, *HLA-C*, *HLA-DRB1* and *HLA-DQB1*) with the phenotypes derived from our health history survey. *HLA* data included the first two fields of the allele name as described in the *HLA* nomenclature, which defines *HLA* allotypes. We limited our analysis to phenotypes reported by a minimum of 100 individuals (case group) and only *HLA* alleles with a frequency greater than 1% (stratified by ancestry) were included in the analysis. *HLA* association testing was performed through logistic regression stratified by self-reported ancestry adjusting for age and sex - except pregnancy condition outcomes or other sex-related phenotypes, which only include age as a covariate. Statistical significance corrected for multiple comparisons was assigned as L ≤ 1.0E-5. For novel associations, conditional analysis was performed when two or more alleles were associated to the same phenotype in order to assess if the alleles were independently associated to the phenotype or if one of the alleles was associated due to linkage disequilibrium. If one previously associated allele lost significance after conditional analysis, it was ruled out of the results. All analyses were conducted using the stats package (version 3.6.2), a component of the R statistical computing environment (R Core Team, 2021; R version 4.3.0, RStudio 2025.05.1 Build 513)(28). A chord diagram was drawn using package circlize(29) (version 0.4.16). To quantify any systematic bias in the PheWAS analysis, we assessed the genomic inflation factor (λ) through chi-square test also in RStudio(28) (2025.05.1 build 513). Quantile-quantile (QQ) plot was generated using package ggplot2(30) (version 3.5.2).

For the replication (AoU) cohort, only the novel associations found in the discovery cohort were analyzed. The logistic regression, stratified by self-reported ancestry, was adjusted by age at last EHR record, sex, first 15 genetic PCs and EHR length in time. The analysis included alleles with a frequency greater than 0.1% with at least 30 individuals carrying the allele of interest. As the replication test was set for the effect in the same direction (i.e., either risk or protection only, one-sided), the statistical significance was assigned as L ≤ 0.1.

For phenotypes for which we detected novel *HLA* associations, we also conducted phenotype-phenotype association analysis. We performed logistic regression adjusting for sex, age and genetic ancestry (“race”) with all other phenotypes as predictors. Only phenotypes reported by a minimum of 100 individuals were included in the analysis. Statistical significance corrected for multiple comparisons was assigned as □ ≤ 1.0E-5. A network diagram was drawn using package ggraph(31) (version 2.2.2). The analysis was conducted also in RStudio(28) (RStudio 2025.05.1 Build 513).

### Molecular docking prediction

The molecular docking prediction for antibiotics and *HLA* Class II was performed through Boltz-2(32) (version: 2.2) in ChimeraX(33) (version: 1.11.dev202510120519). *HLA* Class II sequences were obtained on IPD-IMGT/HLA(34) (Release 3.62) and antibiotics SMILES strings were obtained on PubChem(35) repository. Predictions were performed under default settings.

## Results

### Self-reported health history reveals numerous known and novel HLA-phenotype associations

We identified a total of 48 associations under a significance threshold of 1.0E-5 (45 in EUR, two in API and one in HIS ancestry – no association was found in the AFA ancestry) involving 27 unique alleles and 15 distinct phenotypes in the discovery (NMDP) cohort (Supplementary Table 1). Out of the 48 associations, 15 novel – not previously reported – associations were found (eight as risk), comprising 13 unique alleles and nine distinct phenotypes (Table 2, Figure 1). The novel associations found in the discovery cohort were then analyzed in the replication (AoU) cohort, where 13 out the 15 novel associations were assessed – only cefaclor hypersensitivity and scarlet fever phenotypes were not available for analysis in the AoU cohort. Seven associations were replicated under a p-value ≤ 0.05 (Table 2). The remaining associations were replicated either in a closely related phenotype or in a different ancestry than in the discovery cohort, the majority presented a p-value ≤ 0.1 (Supplementary Table 2). No substantial dose effects were observed (Supplementary Table 3). In addition to clear replication of nearly all novel associations, many associations found in the present study reproduce previous studies’ findings, supporting the validity of the approach and results.

**Figure 1.**
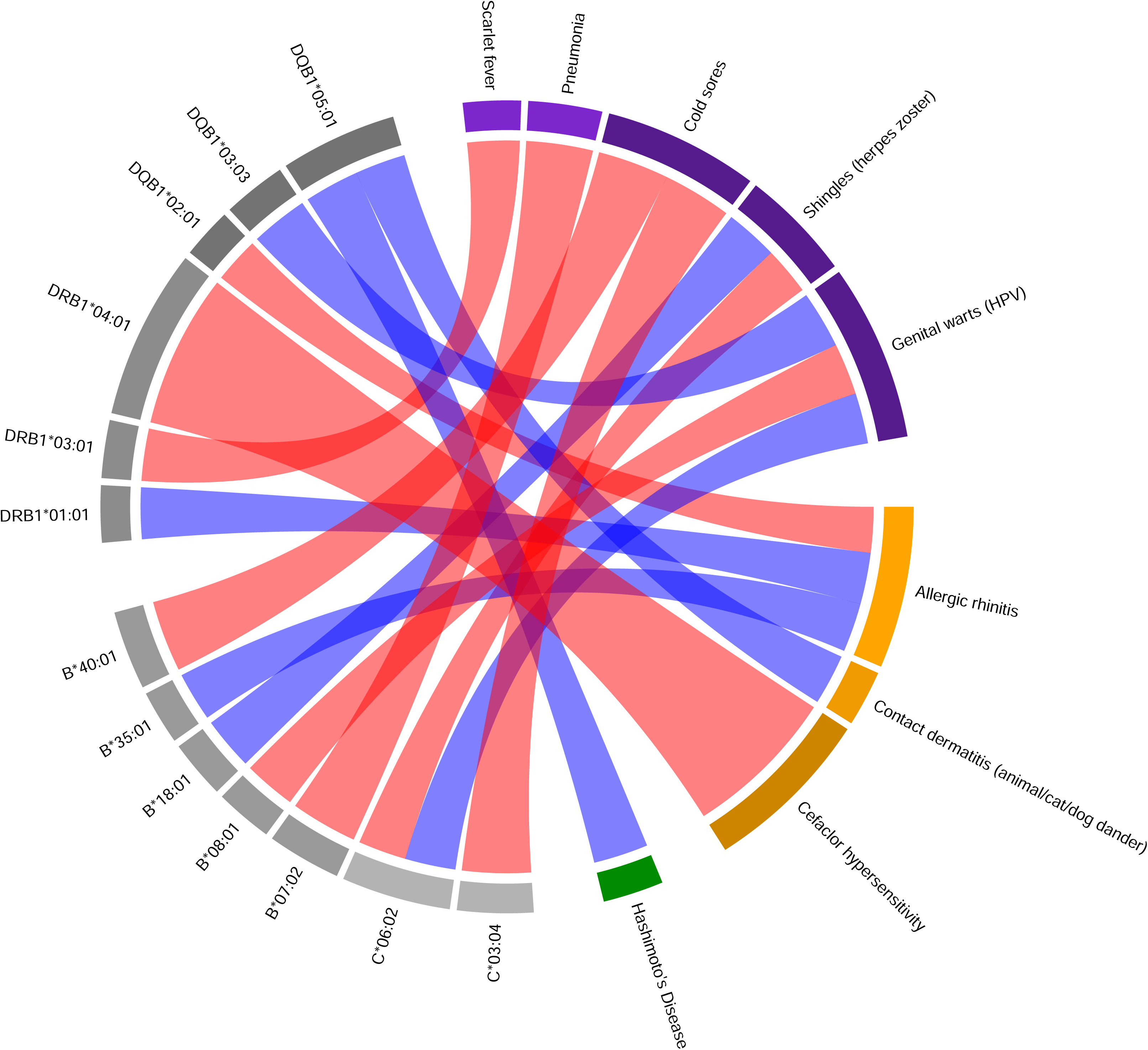
Circle diagram showing novel *HLA*-phenotype associations. Red links: risk association; Blue links: protection association; Grey: *HLA* alleles; Purple: infectious phenotypes; Orange: allergy/hypersensitivity phenotypes; Green: autoimmune phenotypes.

**Table 2.** Novel *HLA* associations in the PheWAS.

| *HLA* allele | Phenotype | Ancestry | NMDP |  |  | All of Us |  |  |
| --- | --- | --- | --- | --- | --- | --- | --- | --- |
|  |  |  | OR | CI<sub>97.5%</sub> | p-value* | OR | CI | p-value** |
| *DRB1\*04:01* | Cefaclor hypersensitivity | EUR | 3.74 | (2.95 - 4.72) | 5.10E-28 | - | - | - |
| *B\*40:01* | Cold sores | API | 1.73 | (1.39 - 2.15) | 7.80E-07 | - | - | - |
| *C\*03:04* | Cold sores | API | 1.67 | (1.34 - 2.07) | 3.70E-06 | - | - | - |
| *B\*07:02* | Pneumonia | HIS | 1.62 | (1.34 - 1.94) | 2.70E-07 | - | - | - |
| *DRB1\*03:01* | Scarlet fever (*S. pyogenes*) | EUR | 1.27 | (1.15 - 1.4) | 2.40E-06 | - | - | - |
| *B\*08:01* | Genital warts (HPV) | EUR | 1.26 | (1.16 - 1.35) | 6.80E-09 | 1.48 | (1.30 - 1.68) | 1.32E-09 |
| *C\*06:02* | Shingles (herpes zoster - VZV) | EUR | 1.17 | (1.09 - 1.25) | 8.90E-06 | - | - | - |
| *DQB1\*02:01* | Allergic rhinitis | EUR | 1.11 | (1.07 - 1.16) | 5.40E-07 | 1.05 | (1.01 - 1.09) | 1.05E-02 |
| *B\*35:01* | Allergic rhinitis | EUR | 0.84 | (0.78 - 0.9) | 6.60E-07 | - | - | - |
| *DQB1\*05:01* | Contact dermatitis (animal/cat/dog dander) | EUR | 0.81 | (0.73 - 0.88) | 4.90E-06 | 0.82 | (0.70 - 0.96) | 1.25E-02 |
| *C\*06:02* | Genital warts (HPV) | EUR | 0.81 | (0.74 - 0.89) | 7.10E-06 | 0.83 | (0.70 - 0.99) | 3.39E-02 |
| *DRB1\*01:01* | Allergic rhinitis | EUR | 0.8 | (0.76 - 0.85) | 3.20E-13 | 0.79 | (0.65 - 0.95) | 1.04E-02 |
| *B\*18:01* | Shingles (herpes zoster - VZV) | EUR | 0.78 | (0.7 - 0.86) | 3.40E-06 | - | - | - |
| *DQB1\*05:01* | Hashimoto's Disease | EUR | 0.75 | (0.66 - 0.85) | 7.90E-06 | 0.81 | (0.74 - 0.88) | 9.64E-07 |
| *DQB1\*03:03* | Genital warts (HPV) | EUR | 0.72 | (0.63 - 0.82) | 1.20E-06 | 0.61 | (0.46 - 0.81) | 4.90E-04 |

OR: odds ratio; CI: confidence interval; \* $\alpha \leq 1.0E-5$ ; \*\* $\alpha \leq 0.05$ .

To detect and quantify systematic biases, we performed a genomic inflation factor test stratified by ancestry and *HLA* locus. The analysis presented a λ ≈ 1 for all five loci and four ancestries, indicating that the test statistics are well-controlled and without bias. Complete genomic inflation factor (λ) values and QQ plots for each loci and ancestry are shown in the Supplementary Figure 1.

Phenotypes for which we detected novel *HLA* associations were also found to co-occur with numerous other conditions. In many cases, related phenotypes frequently co-occurred, but only the index phenotype was characterized by the significant novel *HLA* association. At the same time, HLA associations, both novel and known, are frequently interconnected across multiple phenotypes, underscoring the pleiotropic nature of *HLA* mediated health outcomes. (Supplementary Table 4, Figure 3).

### A novel association with antibiotic hypersensitivity can be attributed to HLA binding specificity

The strongest risk effect identified was the novel association in the EUR ancestry between the common allele *HLA-DRB1*04:01* and hypersensitivity to cefaclor, a beta-lactam antibiotic of the second-generation of cephalosporins(36,37) (OR = 3.74, *p* = 5.10E-28). Questions about specific drug allergy/hypersensitivity were free-text based; thus, responses indicating hypersensitivity or allergy to cefaclor were identified based on two response patterns by participants: the use of “cefaclor”, as the drug active principle name, or “ceclor”, its most common commercial name. Because this phenotype was not available for replication in the AoU cohort, we exploited the two responses as an internal replication by performing a secondary analysis using each term as a separate phenotype. Each was significantly associated with similar effect size (Supplementary Table 5). To explore this further, we analyzed the association with “cephalosporins” (when individuals who reported allergy specifically termed as cephalosporin) and with “cephalosporins class”, comprising all different cephalosporin antibiotics reported in the health history survey (including cefaclor, cefdinir, cefprozil, cefuroxime and cephalexin). Significant associations were found only in the cephalosporins class analysis, suggesting that the link between these *HLA* alleles and cephalosporin allergy is primarily driven by the association with cefaclor (Supplementary Table 5).

To better understand the molecular basis for the *HLA-DRB1*04:01* association with cefaclor hypersensitivity, we compared the molecular docking of *HLA-DRB1*04:01* and cefaclor with another cephalosporin (cephalexin) and two other common antibiotics, penicillin and tetracycline, all non-significant in the *HLA* association analysis. Cephalexin was included in the analysis because it is another cephalosporin presenting a similar number of respondents (245 in EUR ancestry) to cefaclor (287 in EUR ancestry). In turn, Penicillin was selected due the highest number of respondents reporting hypersensitivity/allergy (2943 in EUR), and tetracycline was selected on the basis of showing a similar number of hypersensitivity/allergy respondents (309 in EUR) to cefaclor. In the molecular docking prediction, cefaclor showed values as a strong binder (affinity uM = 1.1) and a binding probability over 50%, while the other three antibiotics presented values as weak binders with lower binding probability (Table 3, Figure 2). Cefaclor is predicted to contact ten amino acids (6 in the b-chain and 4 in the a-chain) in the *HLA-DRB1*04:01* peptide binding groove; cephalexin is predicted to contact eight amino acids (5 in the b-chain and 3 in the a-chain), while tetracycline is predicted to contact seven amino acids (6 in the b-chain and 1 in the a-chain) and penicillin is predicted to contact only three amino acids (2 in the b-chain and 1 in the a-chain). *HLA-DRB1*04:01* and cefaclor binding is predicted to occur in the P4 pocket, where lysine (LYS, pos. 71) and alanine (ALA, pos. 74) are the major anchoring determinants(38) (Figure 2).

**Figure 2.**
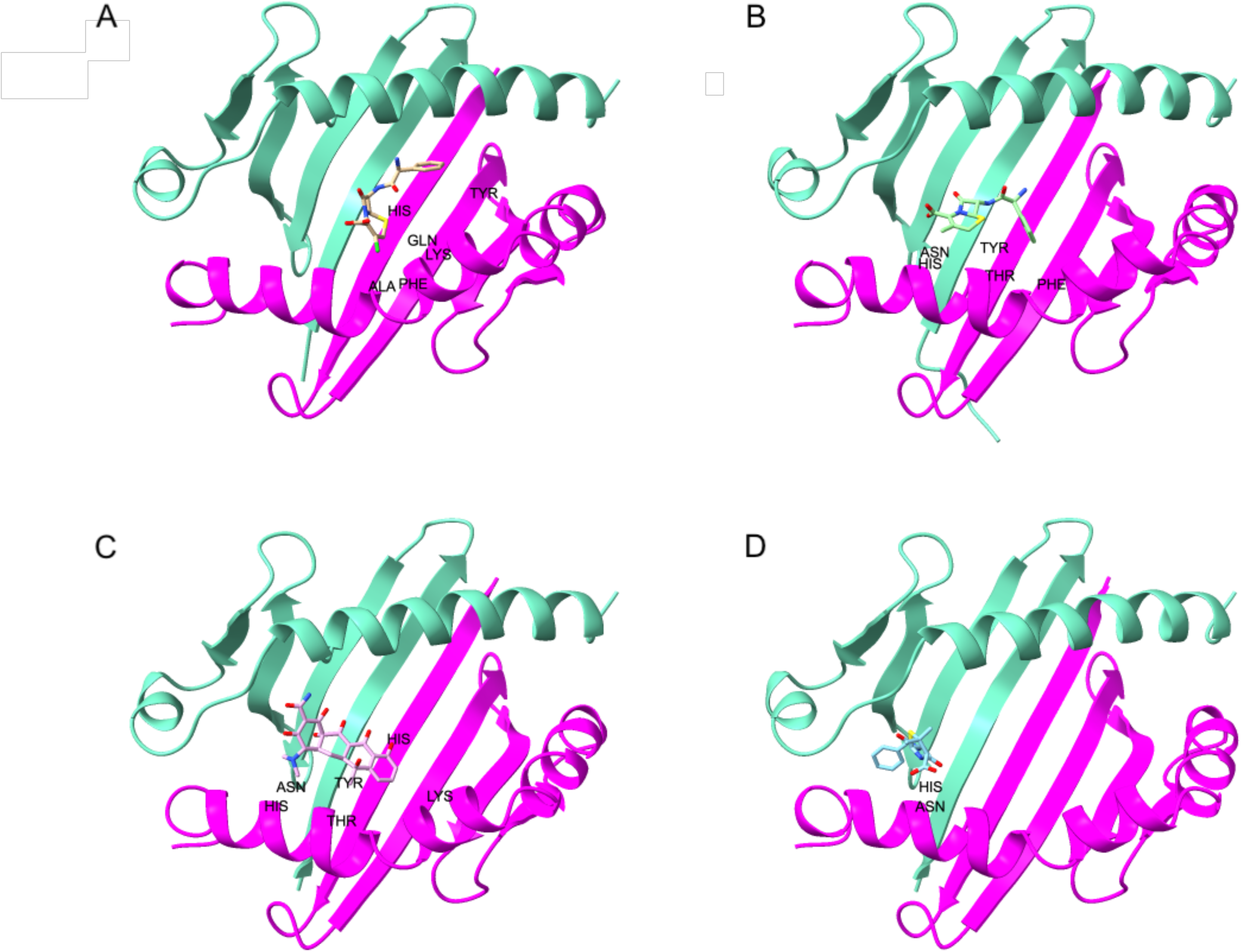
*HLA-DRB1*04:01* Molecular docking prediction. A) Cefaclor. B) Cephalexin. C) Tetracycline. D) Penicillin. L-chain: green; b-chain: magenta. b-chain amino acids binding to the antibiotic molecule are labeled.

**Table 3.** Molecular docking prediction for *HLA-DRB1*04:01* and four common antibiotics.

| Antibiotic | Respondents (N)<sup>†</sup> | Association analysis |  |  |  | Binding prediction |  |  |
| --- | --- | --- | --- | --- | --- | --- | --- | --- |
|  |  | *HLA-DRB1\*04:01* (N) | OR | CI | p-value | ipTM | Affinity (uM) | Binding probability |
| Cefaclor | 287 | 121 | 3.74 | (2.95 - 4.72) | 5.10E-28 | 0.92 | 1.1 | 0.53 |
| Cephalexin | 245 | 51 | 1.34 | (0.97 - 1.81) | 6.40E-02* | 0.83 | 7.6 | 0.12 |
| Tetracycline | 309 | 53 | 1.05 | (0.78 - 1.41) | 7.30E-01* | 0.9 | 5.1 | 0.27 |
| Penicillin | 2943 | 440 | 0.89 | (0.8 - 0.99) | 3.10E-02* | 0.94 | 53 | 0.11 |

<sup>†</sup>EUR ancestry. OR: odds ratio; CI: confidence interval; ipTM: interface predicted template modeling. \*Non-significant.

We also found that cefaclor allergy was strongly associated to augmentin, clavulanate and trimethoprim allergies (OR = 13.44, 12.96 and 10.25, respectively). Drug allergy phenotypes significantly cluster together when considering the associations with strongest effect size (in Figure 3, top ten predictors), although we also observed correlations with other allergy-related phenotypes with smaller effect size (Supplementary Table 4).

**Figure 3.**
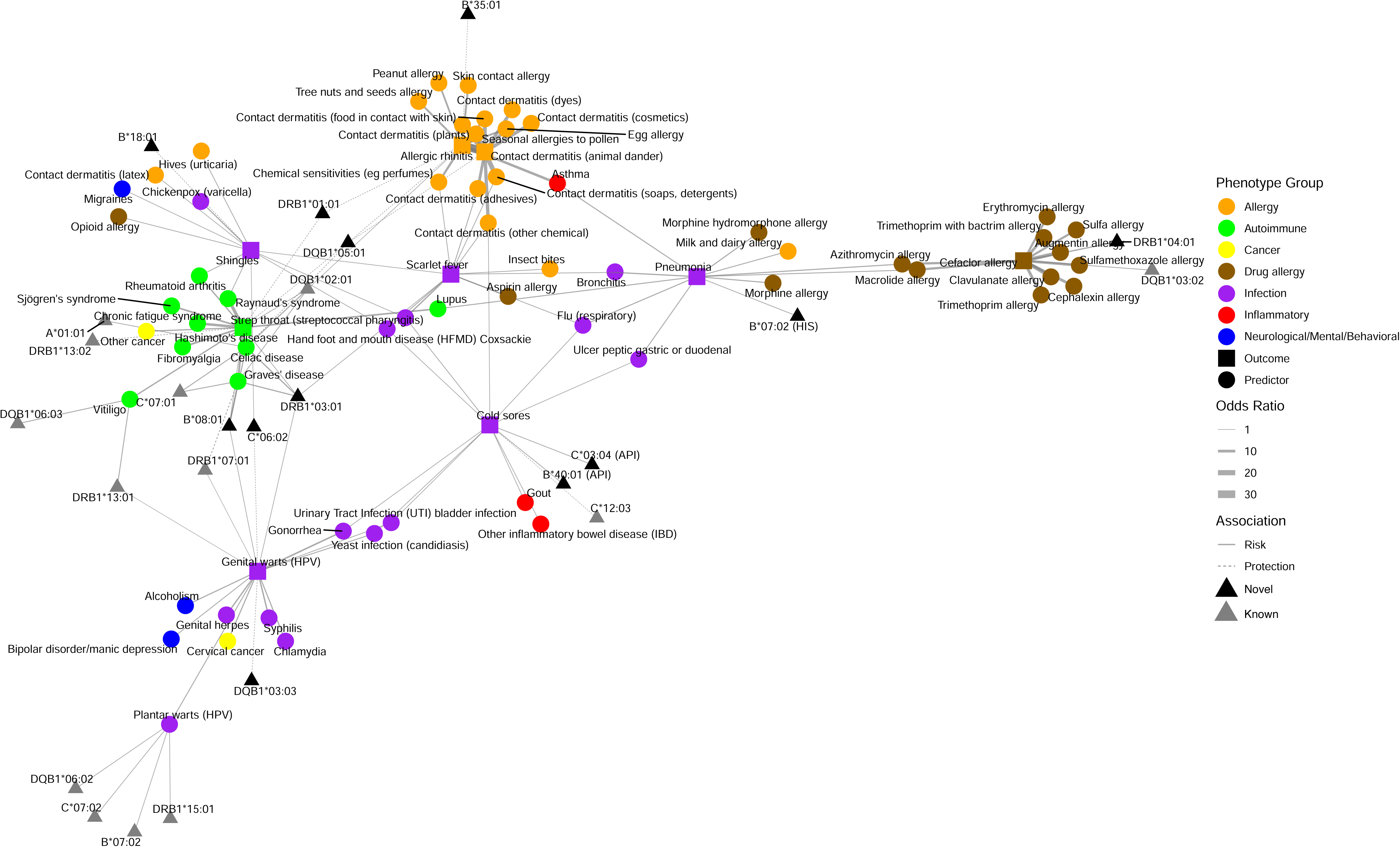
Network diagram with the top 10 phenotype-phenotype associations for phenotypes which we detected novel *HLA* associations.

### Previously unreported HLA associations with common allergies underscore the role of HLA in immune-mediated reactions

In all, allergy/hypersensitivity phenotypes account for five of the novel associations detected in our study, with three distinct phenotypes. Furthering our understanding of the role of *HLA* in allergy, in the EUR population we also identified novel associations between *HLA-DQB1*05:01* (protective) and contact dermatitis to animal dander (OR = 0.81, *p* = 4.90E-06) and between *HLA-DQB1*02:01* (risk, OR = 1.11, *p* = 5.40E-07), *HLA-DRB1*01:01* (protection, OR = 0.8, *p* = 3.20E-13), and *HLA-B*35:01* (protection, OR = 0.84, *p* = 6.60E-07) and allergic rhinitis (Table 2). These results demonstrate that inter-individual variation in allergies and drug hypersensitivities often have immunogenetic underpinnings.

Interestingly, when analyzing the co-occurring allergy-related phenotypes, allergic rhinitis and contact dermatitis (animal dander) present many overlapping relationships (Figure 3). When observing the strongest phenotype associations, allergic rhinitis presented strong relationships with contact dermatitis (animal dander) and pollen allergy (OR = 31.51 and 23.84, respectively), while contact dermatitis (animal dander) was strongly associated to pollen allergy too (OR = 19.39) (Supplementary Table 4). Highlighting a propensity among some individuals to report multiple forms of allergy and airway dysfunction, the significantly correlated phenotypes for both contact dermatitis (animal dander) and allergic rhinitis include asthma, many other contact dermatitis phenotypes and food allergies (egg, peanut and fish, for instance) (Supplementary Table 4).

### Novel associations with viral and bacterial phenotypes emphasize the role of HLA in control of infection

Infectious disease-related phenotypes yielded the highest number of novel associations (nine) in five distinct phenotypes (Table 2). Susceptibility to genital warts presented the most novel associations: *HLA-B*08:01* conferring risk (OR = 1.26, *p* = 6.80E-09) and *HLA-DQB1*03:03* (OR = 0.72, *p* = 1.20E-06) and *HLA-C*06:02* (OR = 0.81, *p* = 7.10E-06) conferring protection in the EUR population.

Demonstrating the role of *HLA* in mediating outcome in infection, *HLA-C*06:02* and *HLA-B*18:01* were associated with risk (OR = 1.17, *p* = 8.90E-06) and protection (OR = 0.78, *p* = 3.40E-06) to shingles, respectively. Shingles, also known as herpes zoster, is a reactivation of the varicella zoster virus (VZV), which remain latent for life subsequent to initial infection(39,40). Interestingly, no associations were found with chickenpox, suggesting that this association is specific to the VZV reactivation rather than primary infection. Likewise, *HLA-DRB1*03:01* was found to confer risk to scarlet fever (OR = 1.27, *p* = 2.40E-06), a manifestation of group A streptococci infection(41,42). In the API population, the *HLA-B*40:01* (OR = 1.27, *p* = 7.80E-07) and *HLA-C*03:04* (OR = 1.67, *p* = 3.70E-06) were associated as risk to cold sores (also known as fever blisters or herpes labialis), caused by the herpes simplex virus (HSV)(43). Finally, in the HIS ancestry, a novel risk association was found between *HLA-B*07:02* and pneumonia (OR = 1.62, *p* = 2.70E-07), although the underlying infectious agent was not reported. These results serve to further underscore the critical role of *HLA* in mediating infection.

We also found that genital warts (HPV) significantly co-occur with other sexually transmitted infections such as gonorrhea (OR = 4.62), syphilis (OR = 4.21) and genital herpes (HSV) (3.71), as well as alcoholism (OR = 3.03) (Figure 3, Supplementary Table 4). As expected, the strongest association with shingles was chickenpox (OR = 3.09) and the strongest association to scarlet fever was strep throat (OR = 3.95); highlighting common transmission, strep throat was also the strongest predictor for cold sores (OR = 1.55). In turn, pneumonia was associated with other respiratory phenotypes such as bronchitis (OR = 3.43), flu (OR = 2.58) and asthma (OR = 2.47), but interestingly also presented associations with drug allergy such as azithromycin and morphine (OR = 2.88 and 2.85, respectively) and the autoimmune disease lupus (OR = 2.71) (Supplementary Table 4).

### Known and novel associations support the pivotal role of HLA in autoimmune disease

*HLA* association with numerous autoimmune conditions has long been recognized. Here, despite examination of a cohort screened at enrollment for immune-mediated disease, a total of 24 associations were found with six autoimmune phenotypes. Well-documented associations, such as *HLA-DRB1*03:01* conferring risk to celiac disease(44) (OR = 3.57, *p* = 4.20E-65) and Graves’ disease(45) (OR = 3.09, *p* = 4.10E-22), and *HLA-B*52:01* as risk to ulcerative colitis(46) (OR = 3.28, *p* = 5.90E-07) were the strongest known associations reproduced in this study. The previously described association between *HLA-DRB1*07:01* and Graves’ disease(47,48) (OR = 0.42, *p* = 7.90E-07) showed the strongest protective effect in the present study (Supplementary Table 1).

At the same time, we detected a novel protective association between *HLA-DQB1*05:01* and Hashimoto’s disease (OR = 0.75, *p* = 7.90E-06, Table 2), demonstrating further the wide-ranging impact of *HLA* variation in autoimmune disease. Strikingly, Hashimoto’s disease was found to frequently co-occur with other autoimmune (or putative autoimmune) conditions, such as Raynaud’s syndrome (OR = 11.26), chronic fatigue syndrome (OR = 8.65) and Graves’ disease (OR = 6.66).

## Discussion

Large *HLA* association studies often face the barriers of sufficient sample size and the high cost for high-resolution *HLA* genotyping. While PheWAS studies utilizing EHR and imputed *HLA* alleles(11,14) have provided insight into *HLA* variation in human health, many common medical conditions remain understudied with respect to immunogenetic variation. We conducted a well-powered examination of high-resolution *HLA* and a wide range of phenotypes encompassing common medical conditions, often under-reported in EHR(18–22), replicated in a large independent cohort. In total, our study resulted in 48 *HLA*-phenotype associations. Our findings corroborate several known associations and uncovers several novel associations, providing new insights into the genetic basis of common health conditions and the high frequency of correlations among them.

The majority of the novel associations are related to phenotypes driven by infectious agents and allergic/hypersensitivity reactions. The most significant and strongest effect among these is the association between *HLA-DRB1*04:01* for cefaclor hypersensitivity in EUR ancestry. To the best of our knowledge, this is the first time that the *HLA-DRB1*04:01* allele, which is relatively common among individuals with European ancestry(49), has been reported as risk for cefaclor hypersensitivity. Cefaclor is a second-generation cephalosporin, which are a group of beta-lactam antibiotics(36,37). Cephalosporins accounts for 6.1% of the documented drug induced anaphylaxis in US, and cefaclor solely is responsible for 0.8%(50). Cefaclor hypersensitivity symptoms vary from mild, such as skin rashes and fever, to more severe and systemic reactions like life-threatening anaphylaxis(36). *HLA* association with beta-lactam antibiotics have been reported(51), but studies in the United States population are scarce. There is no data regarding cefaclor hypersensitivity in the AoU database, which is based on EHR. Discrepancies between patient self-reported and EHR reports of drug allergies and adverse reactions have been reported(52,53), and different recording systems can lead to inconsistencies and discrepancies regarding drug allergy documentation(54,55). Molecular docking prediction showed strong affinity between *HLA-DRB1*04:01* and cefaclor, with the binding occurring in the P4 pocket, supporting the association found. Detection of this strong and robust association with a commonly used antibiotic highlights the need for approaches beyond examination of the EHR to better understand the role of genetic variation in health outcomes.

Further demonstrating the value of our self-report approach, we uncovered three previously undetected associations with genital warts. Caused by the human papillomavirus (HPV), genital (or anogenital) warts is the most frequent sexually transmitted viral infection in the world, manifesting visible lesions as single or multiple papules in both male and female genital organs(56). A systematic review showed that higher rates of anogenital warts are mostly self-reported or from routine genital examinations, suggesting that this condition is likely under-reported in the EHR(18). Similarly, novel associations related to shingles (herpes zoster) and cold sores (herpes simplex) may reflect the fact that these conditions are also often under-reported in EHR. For example, herpes cold sores are frequently occurring, but most episodes are mild and self-treated with over-the-counter topical medications, not generating a medical record(57–59).

Moreover, the average age of our discovery cohort at the time of survey was 39 years; it is highly probable that some health conditions reported in our survey occurred in the respondents’ health histories prior to the widespread use of EHR(60–63). For example, fewer than 20% of physicians reported using even a basic EHR in the early 2000s(64). Policies as the creation of the Office of the National Coordinator for Health Information Technology (ONC)(65) in 2004 and the Health Information Technology for Economic and Clinical Health (HITECH)(61) Act in 2009 later led to rapid uptake: the use of basic EHR in U.S. non-federal hospitals increased from about 9% in 2008 to over 90% by 2015-2017(62). The average age of respondents of the health history survey was 23 years old in 2008, when the use of EHR was minimal. Thus, substantial information about health history in our cohort is likely not reflected in EHR.

Our results also highlight the pleiotropic nature of *HLA* associations with disease. For example, *HLA-C*06:02*, well known as the main risk *HLA* allele for psoriasis(66), was found in two associations not described before: shingles (risk) and genital warts (protection). An interesting interplay between shingles and psoriasis has been observed. Several cases of herpes zoster-induced psoriasis have been reported in the literature(67–69) and an increased risk of psoriasis in patients diagnosed with herpes zoster was demonstrated(70). At the same time, psoriasis itself also showed an increased risk for its patients to develop VZV reactivation(71). The protective association between *HLA-C*06:02* and genital warts adds more complexity to the relationship between this allele and cutaneous conditions with a viral basis.

Our phenotype-phenotype analysis revealed that many allergy-related phenotypes co-occur, specially contact dermatitis and allergic rhinitis, while drug allergy tend to cluster together. In turn, the strongest phenotypes associated to genital warts (HPV) are other sexually transmitted infections, which is related not only to the host-pathogen interactions(72,73), but risk behavior(74,75) and substance use such as alcohol(76) plays a role as well. Interestingly, despite strong associations between many phenotypes, we did not observe consistent sharing of *HLA* risk or protection among these co-occurring conditions. One exception was Hashimoto’s disease, which showed strong associations with other autoimmune conditions. The co-occurrence of these autoimmune phenotypes could be explained by the interplay of *HLA* risk alleles. For instance, *HLA-DRB1*03:01* confers risk to Hashimoto’s disease(77), Graves’ disease(78) and Raynaud’s syndrome(79) or additional shared *HLA* risk alleles(80,81).

Some limitations apply to this work. Our discovery cohort is composed of voluntary bone marrow donors registered in the NMDP database. It is important to highlight some intrinsic characteristics of NMDP registry population: registered donors are, on average, younger, healthier, and have higher socioeconomic status than the general population. Likewise, inconsistency in self-reporting health data cannot be ruled out. However, we emphasize that most of the novel associations reported here were replicated in an independent cohort (AoU), with different participants recruitment processes and demographics. While the in the discovery (NMDP) cohort all phenotypes were collected through self-report questionnaires designed to gather common and often under-reported health data, in the AoU cohort data was obtained from EHR. It is important to note that while several novel associations reported here replicated at nominal significance levels in AoU, they would not have met the significance threshold for phenome-wide discovery in that cohort, and thus would have been overlooked; lower levels of reporting of these conditions in the EHR likely reduces power to detect these associations.

While some phenotypes vary with respect to nomenclature in these cohorts, in most cases we were able to identify reasonable proxies for replication purposes. Genital warts associations – labeled as “anogenital (venereal) warts” in AoU cohort – were replicated, where *HLA-B*08:01* and *HLA-C*07:01* showed risk, while *HLA-C*06:02* and *HLA-DQB1*03:03* showed protection in EUR ancestry. In both cohorts, the association between *HLA-DQB1*03:03* allele and genital warts presented the strongest protective effect. The protective association between *HLA-B*18:01* and shingles (labeled as “herpes zoster”) was replicated as well. However, while *HLA-C*06:02* was found to confer risk to shingles in the discovery cohort, in AoU cohort *HLA-C*06:02* was associated with risk to herpes simplex (HSV), a close relative to varicella-zoster virus (VZV), the infectious agent for shingles. *HLA-C*03:04* was associated with herpes zoster in EUR ancestry, while in the discovery cohort was associated with cold sores in API ancestry. *HLA-B*40:01* association was replicated with HSV (the causal agent for cold sores), but in the EUR ancestry.

In summary, we present robust *HLA* associations across numerous phenotypes, reproducing known associations and identifying novel ones, that were replicated in a large independent cohort. The present study reveals novel high-resolution *HLA* associations based on self-reported health history focused on common conditions that may be under-reported in EHR. These results highlight the complex nature of *HLA* genetic associations and offer new insights into the genetic basis of various health conditions and their implications for personalized medicine, especially in drug allergy related phenotypes.

## Acknowledgements

The authors wish to thank the volunteer donors in the NMDP registry who consent to participate in the health history survey.

## Author contributions

J.A.H. and M.M. conceived this work; J.A.B. and J.A.H. undertook formal analysis, investigation and wrote the original draft; M.M. and M.W. undertook the NMDP dataset collection and curation; J.A.B, S.Y-T.L, R.S., K.G. and A.B. undertook the statistical analysis on the discovery (NMDP) cohort; J.S.H., K.A. and A.A.M-R. undertook the dataset collection and the statistical analysis on the replication (AoU) cohort; J.A.H. obtained resources, conducted project administration and supervised the study; All the authors reviewed and edited the final manuscript.

## Data availability

The full raw data that support the findings of this study are available from the corresponding author upon reasonable request. Data are located in controlled access data storage at the University of California, San Francisco.

## Funding

This work was supported by NIH R01AI158861 (JAH) and [in part] by the Intramural Research Program of the NIH, National Institute of Environmental Health Sciences. The contributions of the NIH author(s) are considered Works of the United States Government. The findings and conclusions presented in this paper are those of the author(s) and do not necessarily reflect the views of the NIH or the U.S. Department of Health and Human Services.

## Competing interests

The authors declare no competing interests.

## Figure legends

**Supplementary Figure 1**. QQ plots for *HLA*-phenotypes associations stratified by ancestry and *HLA* loci. λ = genomic inflation factor; EUR: European descent; AFA: African descent; HIS: Hispanic or Latino; API: Asian or Pacific islander.

## Notes

### Competing Interest Statement

The authors have declared no competing interest.

### Author Declarations

IRB (Institutional Review Board) of University of California, San Francisco gave ethical approval for this work.

