## Supplementary figures and images for "Self-reported health history from 70,724 individuals reveals novel HLA associations with allergy and other frequently underreported conditions"

### Supplementary Figure 1

#### QQ Plot of P−values: HLA−A

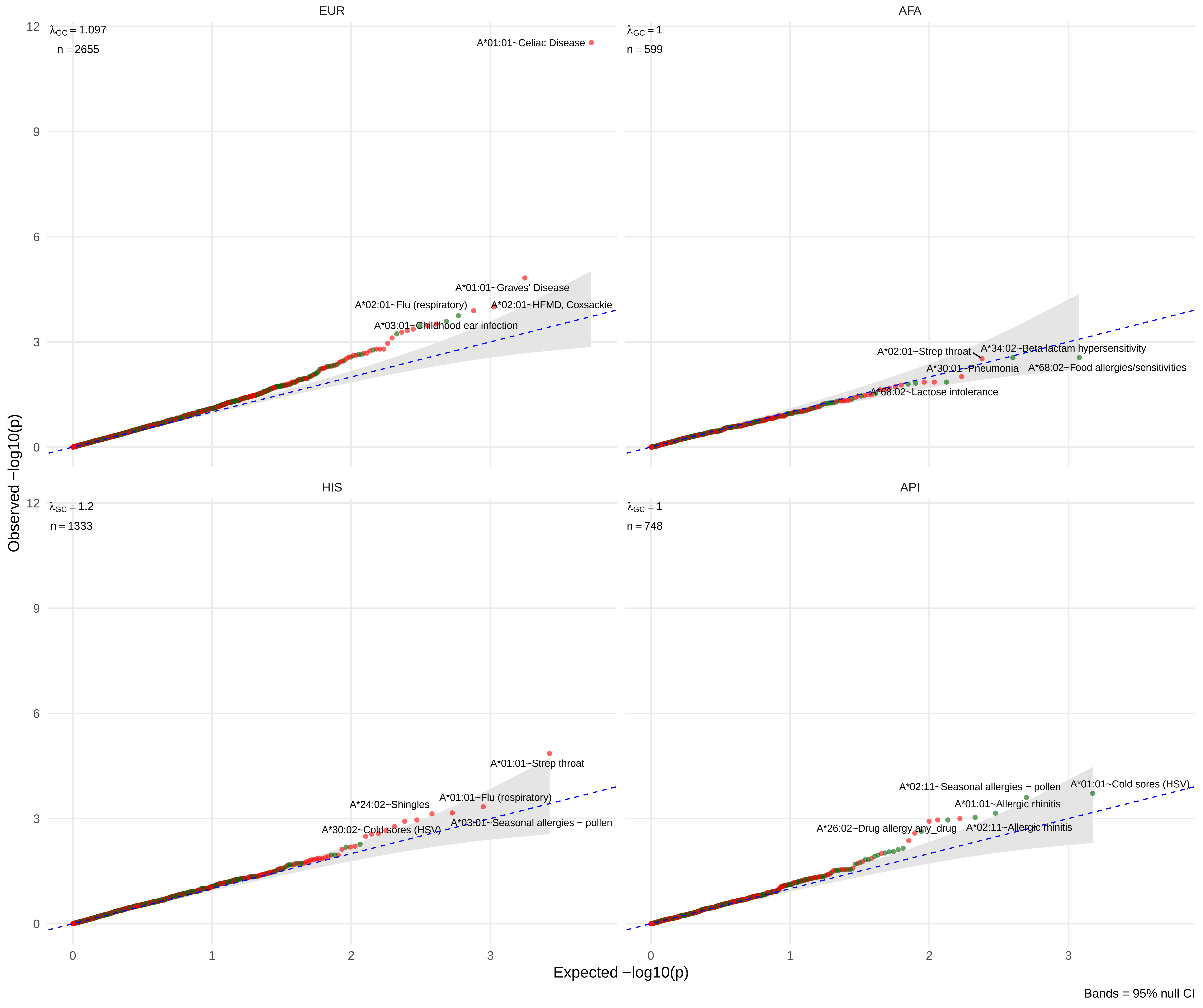

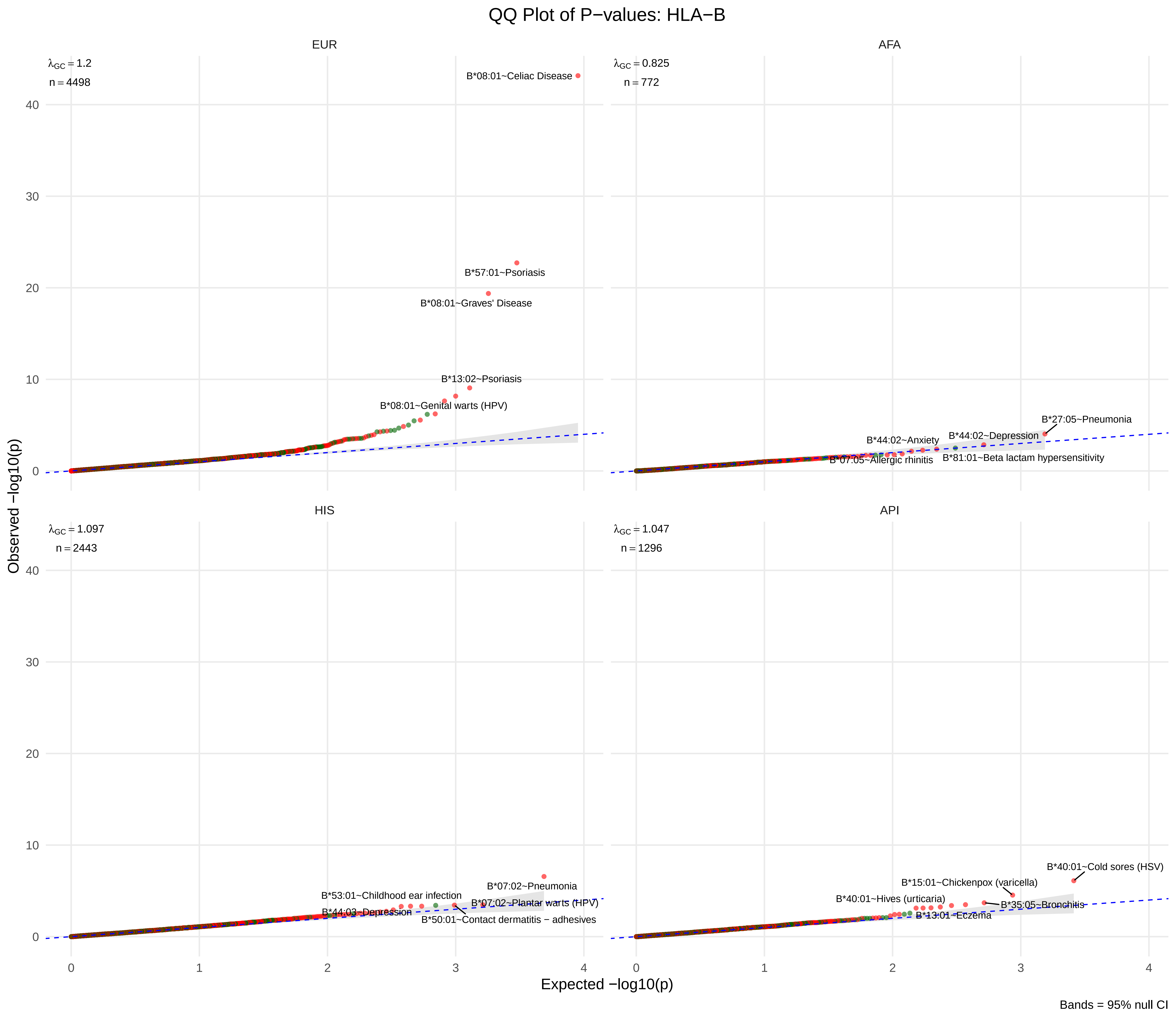

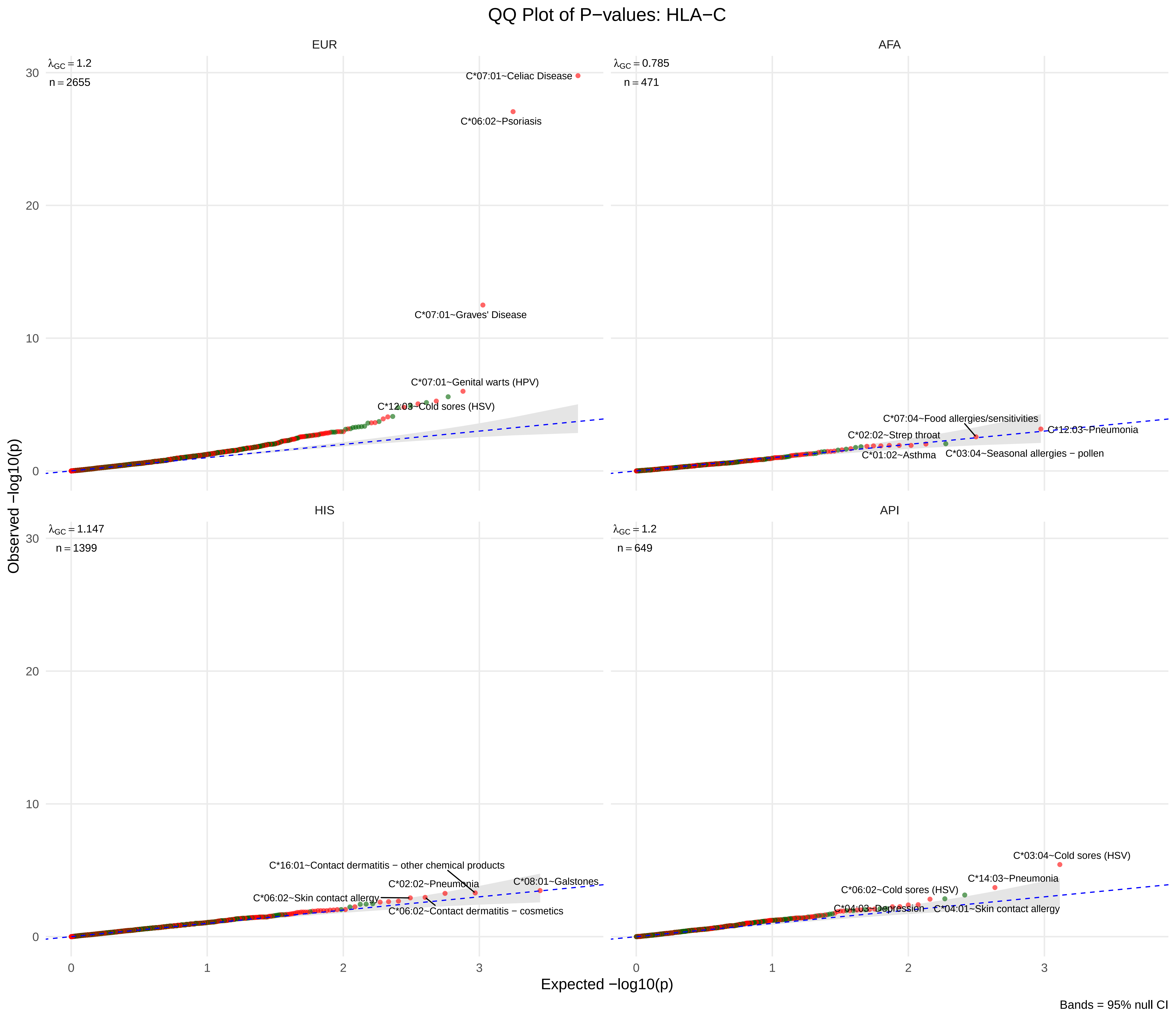

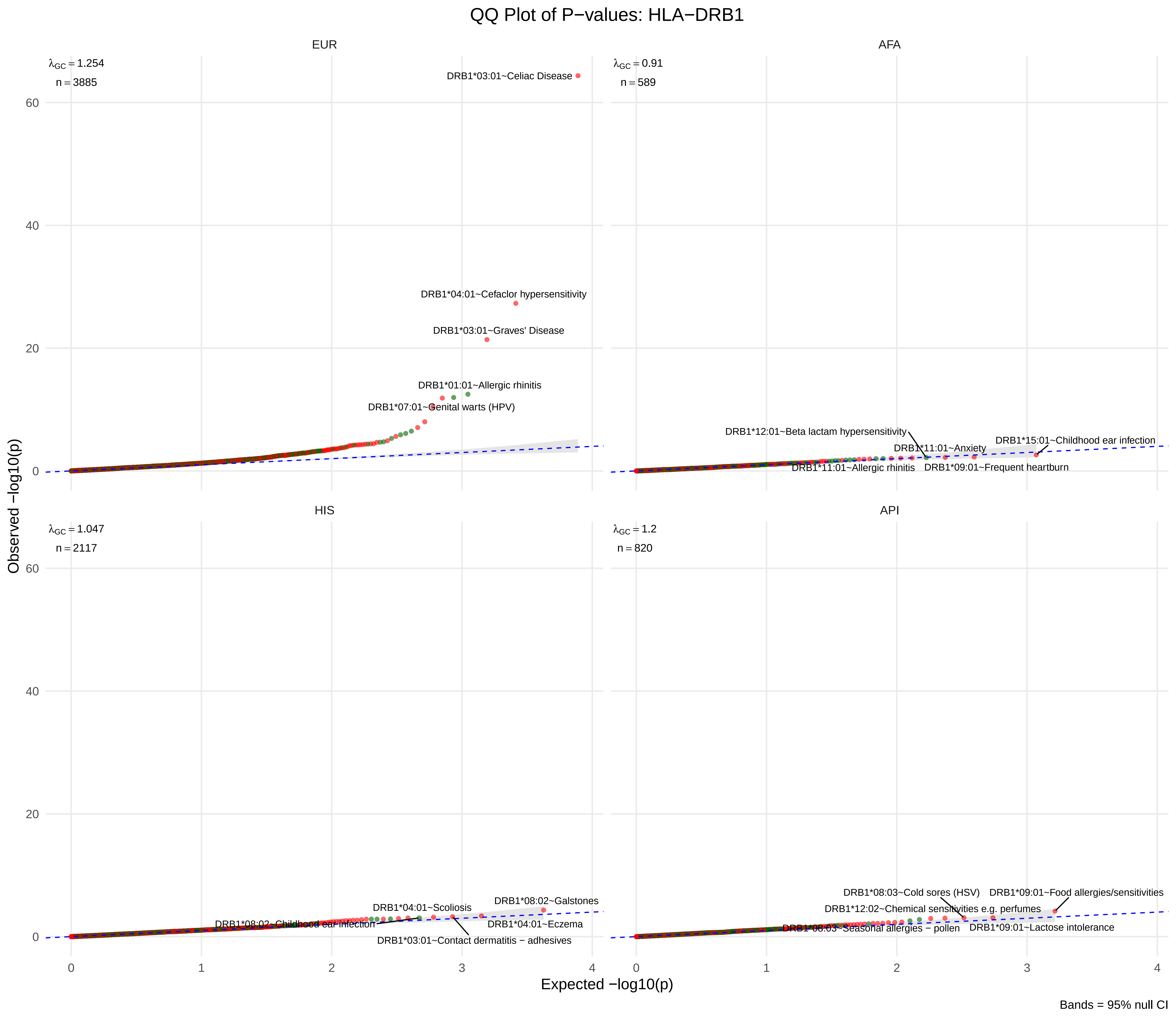

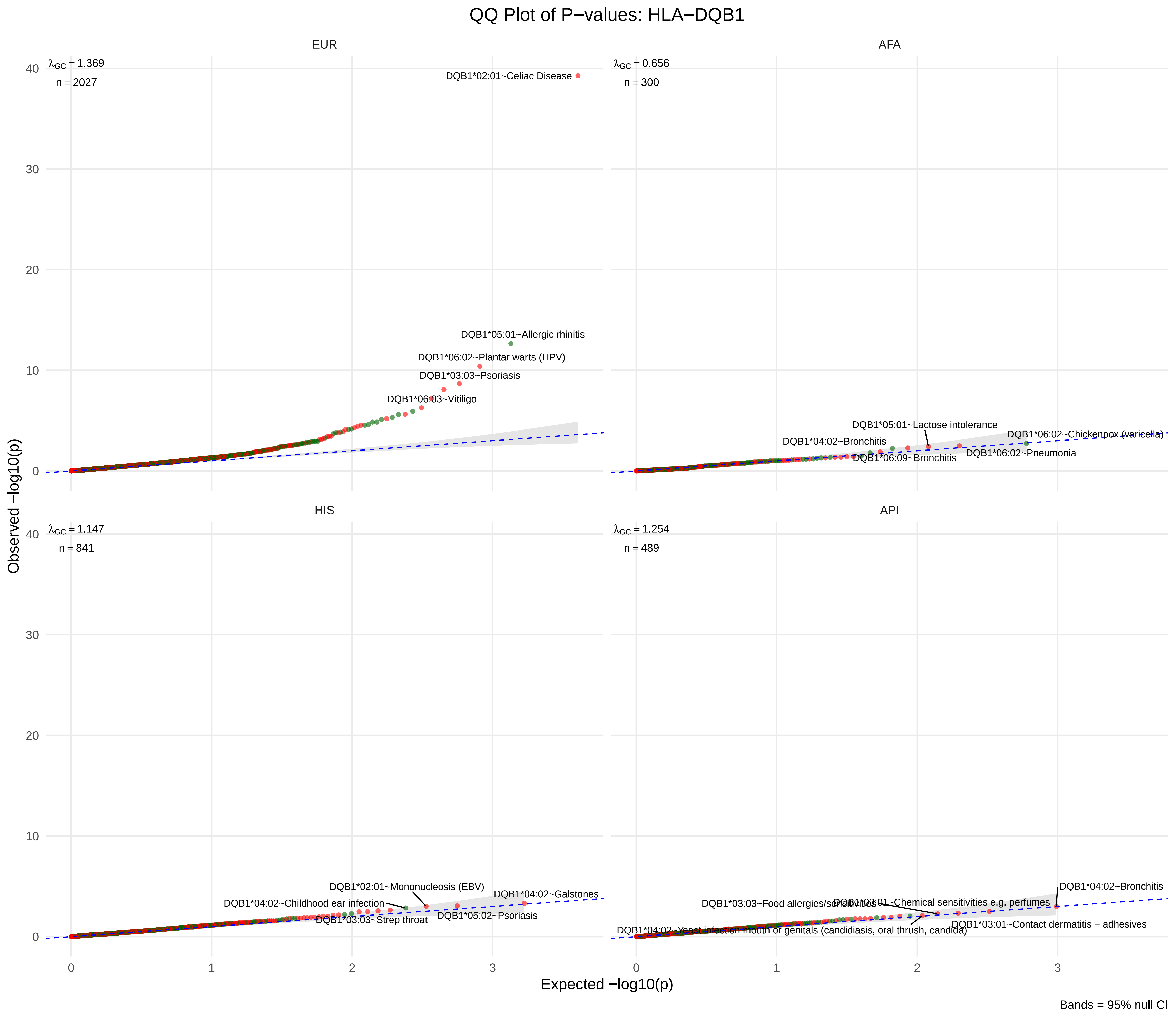
