## Supplementary Table 1 for "Self-reported health history from 70,724 individuals reveals novel HLA associations with allergy and other frequently underreported conditions"

| **Supplementary Table 1.** All significant *HLA* associations in the PheWAS (NMDP cohort). | | | | | | | |
| --- | --- | --- | --- | --- | --- | --- | --- |
| ***HLA* allele** | **Phenotype** | **Ancestry** | **OR** | **CI_97.5%_** | **p-value** | **Case** | **Control** |
| ***DRB1*04:01*** | **Cefaclor hypersensitivity** | **EUR** | **3.74** | **(2.95 - 4.72)** | **5.10E-28** | 121 (0.22) | 10010 (0.09) |
| *DRB1*03:01* | Celiac Disease | EUR | 3.57 | (3.09 - 4.14) | 4.20E-65 | 372 (0.28) | 13809 (0.12) |
| *B*52:01* | Ulcerative colitis | EUR | 3.28 | (1.99 - 5.09) | 5.90E-07 | 19 (0.03) | 1059 (0.01) |
| *DRB1*03:01* | Graves' Disease | EUR | 3.09 | (2.46 - 3.88) | 4.10E-22 | 141 (0.26) | 14040 (0.12) |
| *B*08:01* | Graves' Disease | EUR | 2.94 | (2.33 - 3.7) | 4.20E-20 | 131 (0.24) | 13192 (0.11) |
| *B*08:01* | Celiac Disease | EUR | 2.85 | (2.46 - 3.3) | 6.90E-44 | 317 (0.24) | 13006 (0.11) |
| *DQB1*02:01* | Celiac Disease | EUR | 2.8 | (2.41 - 3.27) | 5.40E-40 | 470 (0.41) | 24003 (0.22) |
| *C*07:01* | Celiac Disease | EUR | 2.36 | (2.03 - 2.73) | 1.70E-30 | 360 (0.28) | 17930 (0.16) |
| *C*07:01* | Graves' Disease | EUR | 2.34 | (1.86 - 2.94) | 3.20E-13 | 146 (0.28) | 18144 (0.16) |
| *DRB1*13:01* | Vitiligo | EUR | 2.32 | (1.72 - 3.06) | 1.00E-08 | 61 (0.12) | 7091 (0.06) |
| *DQB1*06:03* | Vitiligo | EUR | 2.31 | (1.72 - 3.04) | 8.10E-09 | 63 (0.13) | 7393 (0.06) |
| *B*57:01* | Psoriasis | EUR | 2.08 | (1.8 - 2.39) | 1.90E-23 | 226 (0.07) | 4123 (0.03) |
| *DQB1*03:02* | Cefaclor hypersensitivity | EUR | 1.99 | (1.54 - 2.54) | 6.60E-08 | 91 (0.17) | 11668 (0.1) |
| *C*06:02* | Psoriasis | EUR | 1.82 | (1.64 - 2.03) | 8.70E-28 | 479 (0.15) | 10616 (0.09) |
| *B*13:02* | Psoriasis | EUR | 1.76 | (1.46 - 2.09) | 8.60E-10 | 135 (0.04) | 2804 (0.02) |
| ***B*40:01*** | **Cold sores** | **API** | **1.73** | **(1.39 - 2.15)** | **7.80E-07** | 184 (0.11) | 215 (0.07) |
| *A*01:01* | Celiac Disease | EUR | 1.69 | (1.46 - 1.96) | 2.90E-12 | 311 (0.24) | 18666 (0.17) |
| *DQB1*02:01* | Graves' Disease | EUR | 1.69 | (1.35 - 2.13) | 6.50E-06 | 156 (0.3) | 24317 (0.22) |
| ***C*03:04*** | **Cold sores** | **API** | **1.67** | **(1.34 - 2.07)** | **3.70E-06** | 183 (0.11) | 221 (0.07) |
| ***B*07:02*** | **Pneumonia** | **HIS** | **1.62** | **(1.34 - 1.94)** | **2.70E-07** | 186 (0.11) | 573 (0.07) |
| *DQB1*03:03* | Psoriasis | EUR | 1.56 | (1.34 - 1.8) | 2.10E-09 | 218 (0.07) | 5184 (0.04) |
| *B*08:01* | Hashimoto's Disease | EUR | 1.37 | (1.22 - 1.52) | 2.30E-08 | 458 (0.14) | 12865 (0.11) |
| *DRB1*03:01* | Hashimoto's Disease | EUR | 1.34 | (1.2 - 1.49) | 8.60E-08 | 481 (0.16) | 13700 (0.12) |
| *DRB1*03:01* | Genital warts (HPV) | EUR | 1.31 | (1.21 - 1.41) | 1.40E-12 | 1038 (0.15) | 13143 (0.12) |
| *DRB1*15:01* | Plantar warts (HPV) | EUR | 1.3 | (1.2 - 1.4) | 3.60E-11 | 994 (0.16) | 14508 (0.13) |
| *DQB1*06:02* | Plantar warts (HPV) | EUR | 1.3 | (1.2 - 1.4) | 4.10E-11 | 982 (0.16) | 14313 (0.13) |
| ***DRB1*03:01*** | **Scarlet fever (*S. pyogenes*)** | **EUR** | **1.27** | **(1.15 - 1.4)** | **2.40E-06** | 558 (0.15) | 13623 (0.12) |
| *DQB1*02:01* | Hashimoto's Disease | EUR | 1.26 | (1.15 - 1.39) | 2.40E-06 | 770 (0.26) | 23703 (0.22) |
| ***B*08:01*** | **Genital warts (HPV)** | **EUR** | **1.26** | (1.16 - 1.35) | 6.80E-09 | 947 (0.13) | 12376 (0.11) |
| *B*07:02* | Plantar warts (HPV) | EUR | 1.21 | (1.12 - 1.3) | 2.80E-06 | 912 (0.15) | 13979 (0.13) |
| *C*07:02* | Plantar warts (HPV) | EUR | 1.2 | (1.11 - 1.29) | 5.50E-06 | 973 (0.16) | 15060 (0.14) |
| ***C*06:02*** | **Shingles (herpes zoster - VZV)** | **EUR** | **1.17** | **(1.09 - 1.25)** | **8.90E-06** | 1133 (0.11) | 9962 (0.09) |
| ***DQB1*02:01*** | **Allergic rhinitis** | **EUR** | **1.11** | **(1.07 - 1.16)** | **5.40E-07** | 4622 (0.24) | 19851 (0.22) |
| *C*12:03* | Cold sores | EUR | 0.87 | (0.82 - 0.92) | 2.60E-06 | 1674 (0.05) | 4399 (0.05) |
| ***B*35:01*** | **Allergic rhinitis** | **EUR** | **0.84** | **(0.78 - 0.9)** | **6.60E-07** | 1014 (0.05) | 5450 (0.06) |
| *DQB1*05:01* | Allergic rhinitis | EUR | 0.82 | (0.78 - 0.87) | 2.20E-13 | 2141 (0.1) | 11420 (0.12) |
| ***DQB1*05:01*** | **Contact dermatitis (animal/cat/dog dander)** | **EUR** | **0.81** | **(0.73 - 0.88)** | **4.90E-06** | 577 (0.1) | 12984 (0.12) |
| ***C*06:02*** | **Genital warts (HPV)** | **EUR** | **0.81** | **(0.74 - 0.89)** | **7.10E-06** | 575 (0.08) | 10520 (0.1) |
| ***DRB1*01:01*** | **Allergic rhinitis** | **EUR** | **0.8** | **(0.76 - 0.85)** | **3.20E-13** | 1540 (0.07) | 8476 (0.09) |
| ***B*18:01*** | **Shingles (herpes zoster - VZV)** | **EUR** | **0.78** | **(0.7 - 0.86)** | **3.40E-06** | 406 (0.04) | 5049 (0.05) |
| *DRB1*13:01* | Genital warts (HPV) | EUR | 0.77 | (0.68 - 0.86) | 5.10E-06 | 349 (0.05) | 6803 (0.06) |
| ***DQB1*05:01*** | **Hashimoto's Disease** | **EUR** | **0.75** | **(0.66 - 0.85)** | **7.90E-06** | 299 (0.09) | 13262 (0.12) |
| *DRB1*07:01* | Genital warts (HPV) | EUR | 0.74 | (0.68 - 0.81) | 1.10E-12 | 770 (0.11) | 14806 (0.14) |
| ***DQB1*03:03*** | **Genital warts (HPV)** | **EUR** | **0.72** | **(0.63 - 0.82)** | **1.20E-06** | 246 (0.03) | 5156 (0.05) |
| *DRB1*01:01* | Hashimoto's Disease | EUR | 0.68 | (0.58 - 0.78) | 3.30E-07 | 200 (0.06) | 9816 (0.09) |
| *DQB1*05:01* | Celiac Disease | EUR | 0.61 | (0.49 - 0.75) | 2.50E-06 | 107 (0.08) | 13454 (0.12) |
| *DRB1*13:02* | Hashimoto's Disease | EUR | 0.6 | (0.48 - 0.73) | 1.30E-06 | 95 (0.03) | 5368 (0.05) |
| *DRB1*07:01* | Graves' Disease | EUR | 0.42 | (0.29 - 0.58) | 7.90E-07 | 37 (0.07) | 15539 (0.14) |
| Bold: novel associations; OR: odds ratio; CI: confidence interval. | | | | | | | |
