## Supplementary Table 2 for "Self-reported health history from 70,724 individuals reveals novel HLA associations with allergy and other frequently underreported conditions"

| **Supplementary Table 2.** *HLA* associations replicated in the AoU cohort under a different ancestry or closely related phenotype | | | | | |
| --- | --- | --- | --- | --- | --- |
| ***HLA* allele** | **Phenotype** | **Ancestry** | **OR** | **CI** | **p-value** |
| *B*40:01* | Herpes simplex | EUR | 1.08 | (0.99 - 1.18) | 8.12E-02^†^ |
| *C*03:04* | Herpes zoster | EUR | 1.12 | (1.02 - 1.22) | 1.84E-02^‡^ |
| *B*07:02* | Pneumonia | AMR | 1.08 | (0.96 - 1.21) | NS |
| *C*06:02* | Herpes simplex | EUR | 1.07 | (1.00 - 1.15) | 4.53E-02^‡^ |
| *B*35:01* | Allergic rhinitis | AMR | 0.86 | (0.78 - 0.96) | 4.79E-03^‡^ |
| *B*18:01* | Herpes zoster | AMR | 0.72 | (0.51 - 1.02) | 6.40E-02^†^ |
| OR: odds ratio; CI: confidence interval; ^†^ ≤ 0.01; ^‡^ ≤ 0.05. | | | | | |
