## Supplementary Table 3 for "Self-reported health history from 70,724 individuals reveals novel HLA associations with allergy and other frequently underreported conditions"

| **Supplementary Table 3**. Significant associated *HLA* alleles in homozygosis – NMDP cohort | | | | | |
| --- | --- | --- | --- | --- | --- |
|  |  | **Ancestry** | **OR** | **CI_97.5%_** | **p-value*** |
| *DRB1*04:01* | Cefaclor hypersensitivity | EUR | 1.85 | (0.571 - 4.376) | NS |
| *B*40:01* | Cold sores | API | 2.65 | (1.18 - 6.195) | 1.90E-02 |
| *C*03:04* | Cold sores | API | 2.75 | (1.223 - 6.423) | 1.50E-02 |
| *B*07:02* | Pneumonia | HIS | 1.76 | (0.864 - 3.375) | NS |
| *DRB1*03:01* | Scarlet fever (*S. pyogenes*) | EUR | 1.91 | (1.436 - 2.479) | 3.52E-06 |
| *B*08:01* | Genital warts (HPV) | EUR | 1.31 | (0.995 - 1.694) | 4.60E-02 |
| *C*06:02* | Shingles (herpes zoster - VZV) | EUR | 1.22 | (0.924 - 1.584) | NS |
| *DQB1*02:01* | Allergic rhinitis | EUR | 1.19 | (1.089 - 1.304) | 1.22E-04 |
| *B*35:01* | Allergic rhinitis | EUR | 0.61 | (0.382 - 0.936) | 3.20E-02 |
| *DQB1*05:01* | Contact dermatitis (animal/cat/dog dander) | EUR | 0.97 | (0.728 - 1.273) | NS |
| *C*06:02* | Genital warts (HPV) | EUR | 0.59 | (0.37 - 0.895) | 2.00E-02 |
| *DRB1*01:01* | Allergic rhinitis | EUR | 0.59 | (0.437 - 0.79) | 5.47E-04 |
| *B*18:01* | Shingles (herpes zoster - VZV) | EUR | 0.62 | (0.26 - 1.235) | NS |
| *DQB1*05:01* | Hashimoto's Disease | EUR | 0.62 | (0.347 - 1.014) | NS |
| *DQB1*03:03* | Genital warts (HPV) | EUR | 0.52 | (0.161 - 1.255) | NS |
| OR: odds ratio; CI: confidence interval, NS: non-significant; *α ≤ 0.05. | | | | | |
