## Supplementary Table 4 for "Self-reported health history from 70,724 individuals reveals novel HLA associations with allergy and other frequently underreported conditions"

**Supplementary Table 4**. Significant phenotype-phenotype associations

| Phenotype | OR | $CI_{95\%}$ | p-value | N | N (both) |
|---|---|---|---|---|---|
| *Cefaclor allergy* | | | | 291 | |
| Augmentin allergy | 13.44 | (6.99 - 25.82) | 6.4E-15 | 184 | 10 |
| Clavulanate allergy | 12.96 | (6.75 - 24.9) | 1.4E-14 | 190 | 10 |
| Trimethoprim allergy | 10.25 | (6.28 - 16.73) | 1.2E-20 | 446 | 18 |
| Trimethoprim with bactrim allergy | 9.93 | (6.08 - 16.19) | 3.9E-20 | 462 | 18 |
| Cephalexin allergy | 9.88 | (5.18 - 18.87) | 3.8E-12 | 260 | 10 |
| Sulfamethoxazole allergy | 9.73 | (5.97 - 15.87) | 7.9E-20 | 469 | 18 |
| Macrolide allergy | 7.93 | (4.81 - 13.08) | 5.2E-16 | 572 | 17 |
| Azithromycin allergy | 7.80 | (3.62 - 16.8) | 1.5E-07 | 207 | 7 |
| Sulfa allergy | 7.61 | (5.79 - 10.01) | 7.4E-48 | 3097 | 73 |
| Erythromycin allergy | 7.46 | (3.64 - 15.29) | 4.1E-08 | 304 | 8 |
| Amoxicillin allergy | 7.29 | (5.12 - 10.37) | 3.0E-28 | 1267 | 38 |
| Bactrim allergy | 6.28 | (3.3 - 11.93) | 2.0E-08 | 386 | 10 |
| Amoxicillin specific allergy | 6.06 | (4.07 - 9.03) | 6.7E-19 | 1096 | 29 |
| Seasonal allergies to pollen | 5.84 | (4.37 - 7.8) | 5.0E-33 | 28692 | 232 |
| Codeine allergy | 5.60 | (2.95 - 10.65) | 1.5E-07 | 519 | 10 |
| Fruits and vegetables allergy | 4.64 | (2.45 - 8.8) | 2.5E-06 | 557 | 10 |
| Shellfish allergy | 4.26 | (2.32 - 7.83) | 3.1E-06 | 735 | 11 |
| Penicillin allergy | 3.98 | (2.89 - 5.49) | 3.8E-17 | 3228 | 45 |
| Chickenpox (varicella) | 3.29 | (2.32 - 4.67) | 2.3E-11 | 50607 | 247 |
| Contact dermatitis due to adhesives | 3.11 | (2.24 - 4.33) | 1.5E-11 | 3620 | 44 |
| Skin contact allergy | 3.08 | (2.42 - 3.91) | 6.2E-20 | 11975 | 114 |
| Allergic rhinitis | 3.01 | (2.37 - 3.82) | 1.1E-19 | 12779 | 116 |
| Asthma | 2.98 | (2.3 - 3.85) | 1.2E-16 | 8268 | 85 |
| Food allergies or sensitivities | 2.94 | (2.29 - 3.78) | 3.9E-17 | 9671 | 92 |
| Rosacea (acne rosacea) erythematotelangiectatic rosacea (ETR) | 2.92 | (1.86 - 4.58) | 3.1E-06 | 1865 | 21 |
| Contact dermatitis due to soaps or detergents | 2.81 | (2.06 - 3.82) | 4.7E-11 | 4836 | 52 |
| Eczema | 2.72 | (2.05 - 3.61) | 4.4E-12 | 6466 | 63 |
| Hives (urticaria) | 2.61 | (1.86 - 3.66) | 2.8E-08 | 4014 | 40 |
| Contact dermatitis due to animal dander | 2.58 | (1.81 - 3.67) | 1.4E-07 | 3591 | 36 |
| Insect bites | 2.48 | (1.7 - 3.61) | 2.4E-06 | 3323 | 32 |
| Contact dermatitis due to plants (except food) | 2.34 | (1.65 - 3.33) | 2.1E-06 | 4028 | 36 |
| Childhood ear infection | 2.31 | (1.77 - 3.02) | 6.9E-10 | 36629 | 215 |
| Bronchitis | 1.91 | (1.51 - 2.42) | 7.6E-08 | 27855 | 160 |
| Pneumonia | 1.84 | (1.44 - 2.35) | 8.5E-07 | 16127 | 104 |

| | | | | | |
|---|---|---|---|---|---|
| Yeast infection (candidiasis) | 1.84 | (1.42 - 2.38) | 3.8E-06 | 27312 | 153 |
| Mental health/behavioral health condition | 1.79 | (1.4 - 2.27) | 2.2E-06 | 25186 | 155 |
| *Cold sores* | | | | 21453 | |
| Strep throat (streptococcal pharyngitis) | 1.55 | (1.5 - 1.61) | 1.7E-115 | 49317 | 16138 |
| Other inflammatory bowel disease (IBD) indeterminate colitis | 1.50 | (1.27 - 1.78) | 2.7E-06 | 582 | 229 |
| Gonorrhea | 1.47 | (1.26 - 1.73) | 1.6E-06 | 694 | 267 |
| Flu (respiratory) | 1.45 | (1.4 - 1.5) | 2.1E-101 | 41859 | 13995 |
| Gout | 1.44 | (1.24 - 1.67) | 1.6E-06 | 770 | 320 |
| Urinary Tract Infection (UTI) bladder infection | 1.43 | (1.38 - 1.48) | 8.8E-79 | 35934 | 11830 |
| Yeast infection (candidiasis) | 1.43 | (1.38 - 1.48) | 3.4E-81 | 27312 | 9244 |
| Ulcer peptic gastric or duodenal | 1.39 | (1.22 - 1.59) | 1.6E-06 | 946 | 354 |
| Hand foot and mouth disease (HFMD) Coxsackie | 1.39 | (1.3 - 1.48) | 5.9E-22 | 4189 | 1518 |
| Contact dermatitis due to other chemical products | 1.38 | (1.27 - 1.49) | 4.5E-14 | 2627 | 960 |
| Alcoholism | 1.37 | (1.22 - 1.53) | 6.7E-08 | 1338 | 501 |
| Childhood ear infection | 1.35 | (1.3 - 1.39) | 9.8E-69 | 36629 | 12046 |
| Claustrophobia | 1.34 | (1.19 - 1.51) | 1.7E-06 | 1212 | 442 |
| Chickenpox (varicella) | 1.33 | (1.28 - 1.39) | 1.8E-46 | 50607 | 16339 |
| Bee sting | 1.32 | (1.22 - 1.43) | 2.1E-11 | 2683 | 981 |
| Psoriasis | 1.31 | (1.19 - 1.44) | 3.8E-08 | 1920 | 701 |
| Hives (urticaria) | 1.31 | (1.22 - 1.4) | 1.3E-14 | 4014 | 1418 |
| Contact dermatitis due to latex | 1.31 | (1.17 - 1.46) | 1.3E-06 | 1499 | 530 |
| Eczema | 1.30 | (1.23 - 1.38) | 4.0E-21 | 6466 | 2263 |
| Rosacea (acne rosacea) erythematotelangiectatic rosacea (ETR) | 1.30 | (1.18 - 1.43) | 2.2E-07 | 1865 | 669 |
| Contact dermatitis due to soaps or detergents | 1.29 | (1.21 - 1.37) | 1.8E-15 | 4836 | 1692 |
| Bronchitis | 1.29 | (1.24 - 1.33) | 3.4E-49 | 27855 | 9375 |
| Irritable bowel syndrome (IBS) spastic colon | 1.29 | (1.2 - 1.38) | 2.2E-13 | 4164 | 1454 |
| Frequent heartburn | 1.28 | (1.23 - 1.34) | 1.7E-28 | 10600 | 3743 |
| Contact dermatitis due to cosmetics | 1.28 | (1.18 - 1.39) | 8.4E-10 | 3007 | 1043 |
| Plantar warts (HPV) | 1.27 | (1.18 - 1.37) | 6.7E-11 | 3518 | 1244 |
| Chlamydia | 1.27 | (1.18 - 1.36) | 9.8E-11 | 3701 | 1280 |
| Depression | 1.26 | (1.22 - 1.31) | 1.4E-34 | 19092 | 6303 |
| Genital warts (HPV) | 1.26 | (1.18 - 1.35) | 6.4E-12 | 4281 | 1528 |
| Contact dermatitis due to animal dander | 1.26 | (1.17 - 1.35) | 3.3E-10 | 3591 | 1249 |
| Chemical sensitivities (eg perfumes) | 1.24 | (1.17 - 1.32) | 1.5E-11 | 4763 | 1676 |
| Anxiety | 1.24 | (1.19 - 1.29) | 2.2E-29 | 19891 | 6429 |
| Skin contact allergy | 1.24 | (1.18 - 1.29) | 2.7E-22 | 11975 | 4052 |
| Digestive disorder | 1.24 | (1.18 - 1.3) | 1.9E-16 | 7867 | 2700 |
| Lactose intolerance | 1.23 | (1.14 - 1.32) | 1.2E-08 | 3800 | 1290 |

| | | | | | |
|---|---|---|---|---|---|
| Mental health/behavioral health condition | 1.22 | (1.17 - 1.26) | 8.5E-28 | 25186 | 8056 |
| Insect bites | 1.21 | (1.12 - 1.3) | 8.8E-07 | 3323 | 1132 |
| Seasonal allergies to pollen | 1.20 | (1.16 - 1.24) | 4.1E-27 | 28692 | 9377 |
| Contact dermatitis due to adhesives | 1.19 | (1.11 - 1.28) | 1.6E-06 | 3620 | 1217 |
| Food allergies or sensitivities | 1.19 | (1.14 - 1.25) | 2.4E-13 | 9671 | 3224 |
| ADHD/ADD | 1.19 | (1.13 - 1.25) | 5.1E-10 | 7113 | 2303 |
| Allergies | 1.19 | (1.15 - 1.23) | 1.5E-24 | 33879 | 10932 |
| Skin condition | 1.18 | (1.13 - 1.24) | 3.4E-12 | 9651 | 3201 |
| Contact dermatitis due to plants (except food) | 1.17 | (1.1 - 1.26) | 4.4E-06 | 4028 | 1368 |
| Allergic rhinitis | 1.17 | (1.12 - 1.22) | 1.8E-13 | 12779 | 4223 |
| Asthma | 1.17 | (1.11 - 1.23) | 1.2E-09 | 8268 | 2729 |
| Pneumonia | 1.16 | (1.12 - 1.21) | 7.8E-15 | 16127 | 5311 |
| Autoimmune disease | 1.14 | (1.08 - 1.19) | 3.6E-07 | 8733 | 2880 |
| Mononucleosis (EBV) | 1.12 | (1.07 - 1.17) | 2.0E-07 | 12418 | 3964 |
| *Pneumonia* | | | | 16127 | |
| Bronchitis | 3.43 | (3.3 - 3.56) | < 2.2E-16 | 27855 | 10126 |
| Azithromycin allergy | 2.88 | (2.18 - 3.81) | 1.1E-13 | 207 | 97 |
| Morphine allergy | 2.85 | (2.27 - 3.58) | 2.0E-19 | 311 | 148 |
| Morphine hydromorphone allergy | 2.78 | (2.24 - 3.45) | 2.2E-20 | 346 | 162 |
| Lupus | 2.71 | (1.92 - 3.81) | 1.1E-08 | 139 | 63 |
| Flu (respiratory) | 2.58 | (2.48 - 2.69) | < 2.2E-16 | 41859 | 12170 |
| Asthma | 2.47 | (2.35 - 2.59) | 8.4E-282 | 8268 | 3202 |
| Ulcer peptic gastric or duodenal | 2.38 | (2.08 - 2.72) | 1.7E-37 | 946 | 392 |
| Milk and dairy allergy | 2.36 | (1.72 - 3.25) | 1.3E-07 | 163 | 67 |
| Macrolide allergy | 2.35 | (1.99 - 2.79) | 5.9E-23 | 572 | 241 |
| Other inflammatory bowel disease (IBD) indeterminate colitis | 2.33 | (1.97 - 2.76) | 1.2E-22 | 582 | 238 |
| Fibromyalgia | 2.33 | (1.88 - 2.89) | 1.7E-14 | 347 | 149 |
| Cephalosporin specific allergy allergy | 2.33 | (1.63 - 3.31) | 2.8E-06 | 133 | 55 |
| Chronic fatigue syndrome | 2.30 | (1.73 - 3.05) | 1.1E-08 | 202 | 84 |
| Strep throat (streptococcal pharyngitis) | 2.25 | (2.15 - 2.35) | 3.2E-272 | 49317 | 13203 |
| Contact dermatitis due to latex | 2.20 | (1.98 - 2.45) | 2.3E-47 | 1499 | 598 |
| Doxycycline allergy | 2.12 | (1.56 - 2.89) | 1.9E-06 | 175 | 68 |
| Erythromycin allergy | 2.11 | (1.67 - 2.67) | 4.3E-10 | 304 | 121 |
| Contact dermatitis due to dyes | 2.09 | (1.77 - 2.47) | 6.9E-18 | 609 | 233 |
| Hernias | 2.08 | (1.8 - 2.4) | 3.3E-23 | 824 | 319 |
| Fish allergy | 2.06 | (1.53 - 2.78) | 2.1E-06 | 194 | 70 |
| Food additives allergy | 2.06 | (1.51 - 2.81) | 5.8E-06 | 177 | 68 |
| Peanut allergy | 2.03 | (1.61 - 2.55) | 1.3E-09 | 342 | 122 |

| | | | | | |
|---|---|---|---|---|---|
| Opioid allergy | 2.03 | (1.8 - 2.28) | 4.0E-31 | 1206 | 467 |
| Contact dermatitis due to food in contact with the skin | 2.00 | (1.71 - 2.33) | 2.5E-18 | 723 | 271 |
| Cephalexin allergy | 1.95 | (1.51 - 2.52) | 3.9E-07 | 260 | 99 |
| Tree nuts and seeds allergy | 1.95 | (1.63 - 2.33) | 2.0E-13 | 560 | 202 |
| Fluoroquinolone allergy | 1.94 | (1.53 - 2.46) | 3.9E-08 | 306 | 117 |
| Contact dermatitis due to adhesives | 1.94 | (1.8 - 2.08) | 2.7E-72 | 3620 | 1304 |
| Atrial fibrillation | 1.91 | (1.48 - 2.47) | 9.0E-07 | 258 | 94 |
| Migraines | 1.91 | (1.71 - 2.13) | 1.0E-31 | 1518 | 553 |
| Contact dermatitis due to other chemical products | 1.86 | (1.71 - 2.02) | 1.1E-47 | 2627 | 924 |
| Cefaclor allergy | 1.84 | (1.44 - 2.35) | 8.7E-07 | 291 | 104 |
| Appendicitis | 1.84 | (1.56 - 2.17) | 3.4E-13 | 653 | 238 |
| Fruits and vegetables allergy | 1.81 | (1.51 - 2.16) | 1.1E-10 | 557 | 195 |
| Contact dermatitis due to animal dander | 1.79 | (1.67 - 1.93) | 3.9E-55 | 3591 | 1214 |
| Hives (urticaria) | 1.78 | (1.66 - 1.91) | 2.0E-59 | 4014 | 1349 |
| Chemical sensitivities (eg perfumes) | 1.78 | (1.67 - 1.9) | 5.0E-69 | 4763 | 1617 |
| Contact dermatitis due to soaps or detergents | 1.78 | (1.67 - 1.89) | 1.7E-69 | 4836 | 1630 |
| Codeine allergy | 1.78 | (1.48 - 2.13) | 7.8E-10 | 519 | 189 |
| Meningitis | 1.76 | (1.51 - 2.04) | 2.4E-13 | 783 | 273 |
| NSAID (nonsteroidal anti-inflammatory drug) allergy | 1.75 | (1.4 - 2.19) | 1.0E-06 | 361 | 120 |
| Type II Diabetes (diabetes mellitus) | 1.73 | (1.54 - 1.94) | 3.4E-21 | 1452 | 483 |
| Contact dermatitis due to drugs in contact with skin | 1.73 | (1.47 - 2.03) | 4.7E-11 | 666 | 229 |
| Claustrophobia | 1.73 | (1.53 - 1.95) | 2.1E-18 | 1212 | 419 |
| Codeine general allergy | 1.70 | (1.47 - 1.98) | 1.8E-12 | 808 | 282 |
| Osteoarthritis (OA) degenerativejoint disease | 1.70 | (1.57 - 1.85) | 7.6E-37 | 2848 | 995 |
| Bee sting | 1.70 | (1.56 - 1.85) | 1.3E-35 | 2683 | 913 |
| Diahrrhea | 1.70 | (1.59 - 1.8) | 2.3E-62 | 5215 | 1681 |
| Neurological disorder | 1.69 | (1.56 - 1.84) | 7.0E-37 | 2848 | 946 |
| Shellfish allergy | 1.69 | (1.44 - 1.99) | 8.2E-11 | 735 | 237 |
| Childhood ear infection | 1.69 | (1.63 - 1.76) | 2.1E-170 | 36629 | 9970 |
| Mononucleosis (EBV) | 1.69 | (1.62 - 1.77) | 2.2E-122 | 12418 | 3937 |
| Raynaud's syndrome | 1.69 | (1.42 - 2) | 1.7E-09 | 604 | 210 |
| Contact dermatitis due to plants (except food) | 1.68 | (1.57 - 1.8) | 1.9E-48 | 4028 | 1322 |
| Drug allergy | 1.68 | (1.6 - 1.76) | 4.1E-107 | 10734 | 3406 |
| Bipolar disorder/manic depression | 1.67 | (1.51 - 1.85) | 7.2E-23 | 1785 | 583 |
| Gallstones (cholelithiasis) | 1.67 | (1.56 - 1.78) | 1.3E-49 | 4305 | 1444 |
| Insect bites | 1.66 | (1.54 - 1.79) | 2.1E-38 | 3323 | 1074 |
| Rheumatoid arthritis | 1.66 | (1.33 - 2.07) | 8.5E-06 | 360 | 124 |
| Eczema | 1.65 | (1.56 - 1.74) | 2.9E-65 | 6466 | 2008 |

| | | | | | |
|---|---|---|---|---|---|
| Lactose intolerance | 1.64 | (1.52 - 1.76) | 1.9E-40 | 3800 | 1193 |
| Contact dermatitis due to cosmetics | 1.62 | (1.5 - 1.76) | 1.0E-31 | 3007 | 959 |
| Other cardiac arrhythmias (eg SVT) racing of the heart | 1.61 | (1.43 - 1.83) | 3.0E-14 | 1222 | 397 |
| Skin contact allergy | 1.61 | (1.54 - 1.69) | 3.4E-97 | 11975 | 3657 |
| Irritable bowel syndrome (IBS) spastic colon | 1.60 | (1.49 - 1.72) | 1.1E-40 | 4164 | 1332 |
| Food allergies or sensitivities | 1.60 | (1.52 - 1.68) | 9.1E-80 | 9671 | 2939 |
| Celiac disease | 1.59 | (1.37 - 1.85) | 2.3E-09 | 786 | 253 |
| Gluten intolerance (non-celiac) | 1.58 | (1.42 - 1.77) | 6.7E-16 | 1516 | 493 |
| Allergic rhinitis | 1.58 | (1.51 - 1.65) | 4.9E-94 | 12779 | 3815 |
| Contact dermatitis due to metals | 1.56 | (1.44 - 1.69) | 3.2E-26 | 2928 | 909 |
| Penicillin allergy | 1.55 | (1.44 - 1.68) | 1.6E-28 | 3228 | 1020 |
| Allergies | 1.54 | (1.49 - 1.6) | 1.1E-123 | 33879 | 9086 |
| Kidney stones (renal lithiasis) nephrolithiasis | 1.54 | (1.45 - 1.64) | 1.6E-40 | 5020 | 1581 |
| OCD (obsessive complusive disorder) | 1.54 | (1.41 - 1.68) | 2.0E-23 | 2757 | 840 |
| ADHD/ADD | 1.53 | (1.45 - 1.62) | 1.6E-50 | 7113 | 2097 |
| Digestive disorder | 1.53 | (1.45 - 1.62) | 3.6E-57 | 7867 | 2401 |
| Hand foot and mouth disease (HFMD) Coxsackie | 1.53 | (1.43 - 1.64) | 3.1E-33 | 4189 | 1277 |
| Frequent heartburn | 1.53 | (1.46 - 1.6) | 1.5E-70 | 10600 | 3188 |
| Scarlet fever | 1.52 | (1.39 - 1.67) | 8.8E-19 | 2210 | 708 |
| Seasonal allergies to pollen | 1.52 | (1.47 - 1.57) | 1.2E-114 | 28692 | 7844 |
| Urinary Tract Infection (UTI) bladder infection | 1.51 | (1.45 - 1.57) | 2.9E-86 | 35934 | 9441 |
| Plantar warts (HPV) | 1.50 | (1.39 - 1.62) | 5.0E-26 | 3518 | 1091 |
| Anxiety | 1.50 | (1.44 - 1.56) | 1.9E-87 | 19891 | 5502 |
| Yeast infection (candidiasis) | 1.49 | (1.43 - 1.55) | 3.6E-85 | 27312 | 7446 |
| Sulfa allergy | 1.49 | (1.37 - 1.61) | 2.0E-22 | 3097 | 964 |
| Depression | 1.48 | (1.42 - 1.54) | 7.9E-84 | 19092 | 5309 |
| Scoliosis | 1.47 | (1.37 - 1.58) | 8.0E-27 | 4080 | 1226 |
| Mental health/behavioral health condition | 1.47 | (1.42 - 1.53) | 4.3E-89 | 25186 | 6780 |
| Psoriasis | 1.45 | (1.31 - 1.61) | 3.4E-13 | 1920 | 579 |
| Rosacea (acne rosacea) erythematotelangiectatic rosacea (ETR) | 1.44 | (1.3 - 1.6) | 2.0E-12 | 1865 | 570 |
| Heart condition | 1.42 | (1.31 - 1.54) | 4.1E-18 | 3246 | 952 |
| Other digestive | 1.42 | (1.29 - 1.55) | 7.8E-14 | 2382 | 707 |
| Alcoholism | 1.41 | (1.25 - 1.59) | 3.4E-08 | 1338 | 394 |
| Autoimmune disease | 1.39 | (1.32 - 1.46) | 2.2E-36 | 8733 | 2537 |
| Amoxicillin allergy | 1.36 | (1.2 - 1.54) | 1.5E-06 | 1267 | 358 |
| Chickenpox (varicella) | 1.34 | (1.28 - 1.4) | 4.0E-39 | 50607 | 12393 |
| Shingles | 1.32 | (1.24 - 1.4) | 3.3E-20 | 6262 | 1774 |
| Skin condition | 1.28 | (1.22 - 1.35) | 1.5E-22 | 9651 | 2565 |

| | | | | | |
|---|---|---|---|---|---|
| Cold sores | 1.16 | (1.12 - 1.21) | 7.7E-15 | 21453 | 5311 |
| *Scarlet fever* | | | | 2210 | |
| Strep throat (streptococcal pharyngitis) | 3.95 | (3.41 - 4.56) | 4.0E-77 | 49317 | 1993 |
| Aspirin allergy | 3.17 | (1.91 - 5.28) | 8.6E-06 | 176 | 18 |
| Hand foot and mouth disease (HFMD) Coxsackie | 2.17 | (1.89 - 2.5) | 3.5E-28 | 4189 | 251 |
| Contact dermatitis due to adhesives | 1.83 | (1.57 - 2.13) | 3.6E-15 | 3620 | 211 |
| Chemical sensitivities (eg perfumes) | 1.78 | (1.55 - 2.03) | 5.6E-17 | 4763 | 266 |
| Contact dermatitis due to soaps or detergents | 1.66 | (1.45 - 1.91) | 7.4E-13 | 4836 | 250 |
| Flu (respiratory) | 1.66 | (1.51 - 1.83) | 3.6E-26 | 41859 | 1556 |
| Insect bites | 1.65 | (1.39 - 1.94) | 4.5E-09 | 3323 | 168 |
| Contact dermatitis due to other chemical products | 1.64 | (1.36 - 1.97) | 1.3E-07 | 2627 | 133 |
| Bronchitis | 1.63 | (1.5 - 1.78) | 3.5E-28 | 27855 | 1177 |
| Contact dermatitis due to metals | 1.62 | (1.36 - 1.94) | 7.2E-08 | 2928 | 144 |
| Hashimoto's disease | 1.60 | (1.3 - 1.96) | 9.0E-06 | 1854 | 102 |
| Penicillin allergy | 1.60 | (1.35 - 1.88) | 3.2E-08 | 3228 | 163 |
| Contact dermatitis due to cosmetics | 1.59 | (1.33 - 1.89) | 2.2E-07 | 3007 | 151 |
| Scoliosis | 1.59 | (1.36 - 1.85) | 2.4E-09 | 4080 | 199 |
| Mononucleosis (EBV) | 1.58 | (1.43 - 1.75) | 6.8E-20 | 12418 | 591 |
| Plantar warts (HPV) | 1.57 | (1.33 - 1.84) | 4.3E-08 | 3518 | 178 |
| OCD (obsessive complusive disorder) | 1.55 | (1.28 - 1.88) | 8.3E-06 | 2757 | 118 |
| Hives (urticaria) | 1.54 | (1.32 - 1.8) | 4.0E-08 | 4014 | 195 |
| Childhood ear infection | 1.54 | (1.41 - 1.68) | 6.7E-21 | 36629 | 1361 |
| Pneumonia | 1.53 | (1.39 - 1.67) | 5.2E-19 | 16127 | 708 |
| Kidney stones (renal lithiasis) nephrolithiasis | 1.49 | (1.3 - 1.72) | 2.2E-08 | 5020 | 236 |
| Contact dermatitis due to animal dander | 1.49 | (1.26 - 1.76) | 3.2E-06 | 3591 | 161 |
| Skin contact allergy | 1.48 | (1.33 - 1.64) | 9.9E-14 | 11975 | 526 |
| Yeast infection (candidiasis) | 1.47 | (1.34 - 1.61) | 3.6E-16 | 27312 | 1157 |
| Shingles | 1.45 | (1.28 - 1.65) | 1.2E-08 | 6262 | 297 |
| Eczema | 1.45 | (1.27 - 1.65) | 4.7E-08 | 6466 | 273 |
| Drug allergy | 1.43 | (1.28 - 1.59) | 4.7E-11 | 10734 | 475 |
| ADHD/ADD | 1.42 | (1.25 - 1.63) | 2.2E-07 | 7113 | 269 |
| Frequent heartburn | 1.42 | (1.27 - 1.58) | 1.8E-10 | 10600 | 454 |
| Food allergies or sensitivities | 1.42 | (1.27 - 1.59) | 1.5E-09 | 9671 | 400 |
| Anxiety | 1.39 | (1.27 - 1.53) | 9.6E-12 | 19891 | 737 |
| Chickenpox (varicella) | 1.39 | (1.24 - 1.56) | 1.4E-08 | 50607 | 1792 |
| Urinary Tract Infection (UTI) bladder infection | 1.38 | (1.25 - 1.52) | 6.5E-11 | 35934 | 1395 |
| Allergic rhinitis | 1.37 | (1.23 - 1.51) | 3.0E-09 | 12779 | 510 |
| Asthma | 1.34 | (1.19 - 1.52) | 2.1E-06 | 8268 | 326 |

| | | | | | |
|---|---|---|---|---|---|
| Autoimmune disease | 1.34 | (1.2 - 1.51) | 3.8E-07 | 8733 | 395 |
| Allergies | 1.34 | (1.23 - 1.46) | 3.4E-11 | 33879 | 1233 |
| Mental health/behavioral health condition | 1.34 | (1.22 - 1.47) | 2.7E-10 | 25186 | 902 |
| Seasonal allergies to pollen | 1.29 | (1.18 - 1.41) | 6.5E-09 | 28692 | 1056 |
| Depression | 1.25 | (1.13 - 1.37) | 5.4E-06 | 19092 | 680 |
| *Genital warts (HPV)* | | | | 4281 | |
| Gonorrhea | 4.62 | (3.77 - 5.66) | 1.8E-49 | 694 | 129 |
| Syphilis | 4.21 | (2.6 - 6.82) | 5.0E-09 | 130 | 22 |
| Genital herpes | 3.71 | (3.34 - 4.13) | 6.0E-130 | 2494 | 512 |
| Alcoholism | 3.03 | (2.58 - 3.56) | 7.2E-42 | 1338 | 193 |
| Cervical cancer | 2.96 | (1.97 - 4.45) | 1.8E-07 | 147 | 30 |
| Chlamydia | 2.92 | (2.64 - 3.23) | 1.9E-96 | 3701 | 537 |
| Yeast infection (candidiasis) | 2.34 | (2.18 - 2.51) | 1.5E-121 | 27312 | 2616 |
| Plantar warts (HPV) | 2.19 | (1.96 - 2.44) | 4.9E-45 | 3518 | 437 |
| Bipolar disorder/manic depression | 1.89 | (1.6 - 2.24) | 6.1E-14 | 1785 | 166 |
| Urinary Tract Infection (UTI) bladder infection | 1.78 | (1.65 - 1.92) | 3.7E-53 | 35934 | 2892 |
| Chickenpox (varicella) | 1.72 | (1.58 - 1.88) | 4.6E-34 | 50607 | 3607 |
| Mental health/behavioral health condition | 1.71 | (1.6 - 1.82) | 4.3E-56 | 25186 | 1877 |
| Claustrophobia | 1.70 | (1.4 - 2.07) | 7.5E-08 | 1212 | 121 |
| Depression | 1.70 | (1.58 - 1.82) | 7.3E-52 | 19092 | 1497 |
| Anxiety | 1.68 | (1.57 - 1.8) | 3.1E-48 | 19891 | 1496 |
| ADHD/ADD | 1.67 | (1.51 - 1.84) | 4.4E-25 | 7113 | 556 |
| Ulcer peptic gastric or duodenal | 1.64 | (1.32 - 2.04) | 6.5E-06 | 946 | 101 |
| Irritable bowel syndrome (IBS) spastic colon | 1.56 | (1.39 - 1.74) | 3.3E-14 | 4164 | 369 |
| Strep throat (streptococcal pharyngitis) | 1.48 | (1.38 - 1.6) | 1.7E-24 | 49317 | 3287 |
| Rosacea (acne rosacea) erythematotelangiectatic rosacea (ETR) | 1.45 | (1.23 - 1.7) | 6.2E-06 | 1865 | 182 |
| OCD (obsessive complusive disorder) | 1.44 | (1.24 - 1.68) | 2.1E-06 | 2757 | 201 |
| Digestive disorder | 1.42 | (1.3 - 1.56) | 6.1E-15 | 7867 | 657 |
| Insect bites | 1.36 | (1.19 - 1.55) | 5.0E-06 | 3323 | 276 |
| Lactose intolerance | 1.36 | (1.19 - 1.54) | 2.7E-06 | 3800 | 290 |
| Frequent heartburn | 1.34 | (1.24 - 1.46) | 5.1E-13 | 10600 | 838 |
| Flu (respiratory) | 1.32 | (1.24 - 1.42) | 7.9E-17 | 41859 | 2791 |
| Skin condition | 1.28 | (1.18 - 1.4) | 1.1E-08 | 9651 | 716 |
| Cold sores | 1.26 | (1.18 - 1.35) | 9.7E-12 | 21453 | 1528 |
| Mononucleosis (EBV) | 1.25 | (1.16 - 1.35) | 1.6E-08 | 12418 | 933 |
| Bronchitis | 1.25 | (1.17 - 1.33) | 7.6E-12 | 27855 | 2001 |
| Skin contact allergy | 1.23 | (1.14 - 1.33) | 2.2E-07 | 11975 | 883 |
| Drug allergy | 1.23 | (1.14 - 1.34) | 4.4E-07 | 10734 | 832 |

| | | | | | |
|---|---|---|---|---|---|
| Childhood ear infection | 1.23 | (1.15 - 1.31) | 5.1E-10 | 36629 | 2343 |
| Seasonal allergies to pollen | 1.23 | (1.15 - 1.31) | 3.6E-10 | 28692 | 1963 |
| Allergies | 1.22 | (1.15 - 1.3) | 5.7E-10 | 33879 | 2279 |
| Food allergies or sensitivities | 1.22 | (1.12 - 1.33) | 7.1E-06 | 9671 | 689 |
| *Shingles* | | | | 6262 | |
| Chickenpox (varicella) | 3.09 | (2.84 - 3.37) | 1.7E-145 | 50607 | 5609 |
| Rheumatoid arthritis | 1.96 | (1.49 - 2.57) | 1.2E-06 | 360 | 67 |
| Raynaud's syndrome | 1.71 | (1.37 - 2.14) | 2.6E-06 | 604 | 94 |
| Opioid allergy | 1.53 | (1.29 - 1.81) | 7.1E-07 | 1206 | 172 |
| Hives (urticaria) | 1.52 | (1.38 - 1.68) | 1.9E-16 | 4014 | 503 |
| Strep throat (streptococcal pharyngitis) | 1.48 | (1.39 - 1.58) | 1.5E-34 | 49317 | 4773 |
| Hand foot and mouth disease (HFMD) Coxsackie | 1.48 | (1.34 - 1.64) | 3.4E-14 | 4189 | 482 |
| Migraines | 1.47 | (1.25 - 1.72) | 2.1E-06 | 1518 | 185 |
| Contact dermatitis due to latex | 1.46 | (1.25 - 1.72) | 2.7E-06 | 1499 | 184 |
| Scarlet fever | 1.45 | (1.27 - 1.65) | 1.4E-08 | 2210 | 297 |
| Contact dermatitis due to animal dander | 1.41 | (1.27 - 1.58) | 4.4E-10 | 3591 | 406 |
| Contact dermatitis due to other chemical products | 1.41 | (1.25 - 1.6) | 6.9E-08 | 2627 | 308 |
| Yeast infection (candidiasis) | 1.39 | (1.31 - 1.48) | 2.0E-28 | 27312 | 2909 |
| Irritable bowel syndrome (IBS) spastic colon | 1.38 | (1.25 - 1.53) | 3.8E-10 | 4164 | 477 |
| Neurological disorder | 1.37 | (1.22 - 1.55) | 2.2E-07 | 2848 | 331 |
| Flu (respiratory) | 1.36 | (1.28 - 1.43) | 4.2E-27 | 41859 | 4095 |
| Contact dermatitis due to adhesives | 1.35 | (1.21 - 1.51) | 5.9E-08 | 3620 | 416 |
| Asthma | 1.35 | (1.25 - 1.45) | 2.8E-14 | 8268 | 883 |
| Chemical sensitivities (eg perfumes) | 1.34 | (1.22 - 1.48) | 7.6E-10 | 4763 | 561 |
| Sulfa allergy | 1.34 | (1.2 - 1.5) | 2.9E-07 | 3097 | 384 |
| Insect bites | 1.33 | (1.18 - 1.49) | 1.4E-06 | 3323 | 365 |
| Plantar warts (HPV) | 1.33 | (1.19 - 1.48) | 4.0E-07 | 3518 | 412 |
| Pneumonia | 1.32 | (1.25 - 1.41) | 1.6E-20 | 16127 | 1774 |
| Drug allergy | 1.32 | (1.23 - 1.41) | 1.1E-15 | 10734 | 1219 |
| Frequent heartburn | 1.31 | (1.22 - 1.4) | 8.4E-15 | 10600 | 1201 |
| Bronchitis | 1.30 | (1.23 - 1.37) | 4.5E-22 | 27855 | 2928 |
| Mononucleosis (EBV) | 1.29 | (1.21 - 1.38) | 8.1E-15 | 12418 | 1365 |
| Urinary Tract Infection (UTI) bladder infection | 1.29 | (1.22 - 1.37) | 5.2E-17 | 35934 | 3586 |
| Digestive disorder | 1.28 | (1.19 - 1.39) | 3.9E-10 | 7867 | 868 |
| Gallstones (cholelithiasis) | 1.28 | (1.16 - 1.41) | 7.9E-07 | 4305 | 527 |
| Allergic rhinitis | 1.27 | (1.19 - 1.35) | 7.6E-13 | 12779 | 1338 |
| Eczema | 1.26 | (1.16 - 1.38) | 2.0E-07 | 6466 | 662 |
| Autoimmune disease | 1.24 | (1.15 - 1.33) | 9.7E-09 | 8733 | 1008 |

| | | | | | |
|---|---|---|---|---|---|
| Seasonal allergies to pollen | 1.24 | (1.18 - 1.31) | 2.5E-15 | 28692 | 2857 |
| Allergies | 1.23 | (1.16 - 1.29) | 3.3E-14 | 33879 | 3309 |
| ADHD/ADD | 1.23 | (1.12 - 1.34) | 7.4E-06 | 7113 | 641 |
| Food allergies or sensitivities | 1.22 | (1.13 - 1.31) | 1.4E-07 | 9671 | 977 |
| Skin contact allergy | 1.22 | (1.14 - 1.3) | 1.1E-08 | 11975 | 1223 |
| Childhood ear infection | 1.21 | (1.14 - 1.28) | 6.8E-12 | 36629 | 3394 |
| Anxiety | 1.17 | (1.1 - 1.25) | 3.4E-07 | 19891 | 1744 |
| Depression | 1.16 | (1.1 - 1.24) | 9.7E-07 | 19092 | 1724 |
| Mental health/behavioral health condition | 1.16 | (1.09 - 1.23) | 4.9E-07 | 25186 | 2221 |
| *Allergic Rhinitis* | | | | 12779 | |
| Contact dermatitis due to animal dander | 31.51 | (28.68 - 34.63) | < 2.2E-16 | 3591 | 3029 |
| Seasonal allergies to pollen | 23.84 | (22.36 - 25.41) | < 2.2E-16 | 28692 | 11607 |
| Asthma | 8.68 | (8.26 - 9.13) | < 2.2E-16 | 8268 | 4654 |
| Chemical sensitivities (eg perfumes) | 7.69 | (7.22 - 8.19) | < 2.2E-16 | 4763 | 2760 |
| Contact dermatitis due to food in contact with the skin | 7.14 | (6.12 - 8.33) | 1.2E-138 | 723 | 444 |
| Skin contact allergy | 6.59 | (6.31 - 6.9) | < 2.2E-16 | 11975 | 5686 |
| Tree nuts and seeds allergy | 6.31 | (5.31 - 7.5) | 1.5E-97 | 560 | 328 |
| Egg allergy | 6.30 | (4.24 - 9.36) | 9.0E-20 | 104 | 61 |
| Contact dermatitis due to plants (except food) | 6.00 | (5.61 - 6.41) | < 2.2E-16 | 4028 | 2151 |
| Peanut allergy | 5.83 | (4.68 - 7.27) | 9.8E-56 | 342 | 195 |
| Contact dermatitis due to soaps or detergents | 5.72 | (5.38 - 6.08) | < 2.2E-16 | 4836 | 2495 |
| Contact dermatitis due to dyes | 5.38 | (4.56 - 6.33) | 3.6E-90 | 609 | 332 |
| Eczema | 5.36 | (5.08 - 5.67) | < 2.2E-16 | 6466 | 3171 |
| Contact dermatitis due to other chemical products | 5.32 | (4.91 - 5.77) | < 2.2E-16 | 2627 | 1378 |
| Contact dermatitis due to adhesives | 5.28 | (4.92 - 5.66) | < 2.2E-16 | 3620 | 1851 |
| Fish allergy | 5.26 | (3.94 - 7.02) | 2.1E-29 | 194 | 104 |
| Contact dermatitis due to cosmetics | 5.26 | (4.87 - 5.67) | < 2.2E-16 | 3007 | 1557 |
| Contact dermatitis due to drugs in contact with skin | 5.05 | (4.32 - 5.9) | 3.5E-93 | 666 | 353 |
| Food allergies or sensitivities | 4.74 | (4.52 - 4.96) | < 2.2E-16 | 9671 | 4240 |
| Insect bites | 4.73 | (4.4 - 5.09) | < 2.2E-16 | 3323 | 1636 |
| Contact dermatitis due to metals | 4.67 | (4.33 - 5.05) | < 2.2E-16 | 2928 | 1445 |
| Hives (urticaria) | 4.67 | (4.37 - 4.99) | < 2.2E-16 | 4014 | 1941 |
| Contact dermatitis due to latex | 4.46 | (4.01 - 4.95) | 3.5E-172 | 1499 | 735 |
| Fruits and vegetables allergy | 4.44 | (3.75 - 5.27) | 5.8E-66 | 557 | 278 |
| Milk and dairy allergy | 4.44 | (3.25 - 6.06) | 8.5E-21 | 163 | 79 |
| Food additives allergy | 4.20 | (3.11 - 5.68) | 1.0E-20 | 177 | 87 |
| Shellfish allergy | 4.15 | (3.58 - 4.82) | 7.7E-78 | 735 | 349 |
| Lactose intolerance | 4.00 | (3.73 - 4.28) | < 2.2E-16 | 3800 | 1692 |

| | | | | | |
|---|---|---|---|---|---|
| Aspirin allergy | 3.77 | (2.77 - 5.12) | 2.6E-17 | 176 | 83 |
| Clindamycin allergy | 3.55 | (2.53 - 4.99) | 2.9E-13 | 143 | 64 |
| NSAID (nonsteroidal anti-inflammatory drug) allergy | 3.52 | (2.84 - 4.36) | 2.0E-30 | 361 | 162 |
| Fluoroquinolone allergy | 3.50 | (2.78 - 4.41) | 2.6E-26 | 306 | 134 |
| Cephalexin allergy | 3.50 | (2.72 - 4.5) | 1.6E-22 | 260 | 115 |
| Gluten intolerance (non-celiac) | 3.47 | (3.12 - 3.86) | 6.8E-118 | 1516 | 652 |
| Drug allergy | 3.44 | (3.28 - 3.6) | < 2.2E-16 | 10734 | 3973 |
| Bee sting | 3.39 | (3.12 - 3.67) | 8.1E-192 | 2683 | 1102 |
| Ciprofloxacin allergy | 3.27 | (2.43 - 4.4) | 5.7E-15 | 185 | 79 |
| Alopecia (alopecia areata) spot baldness | 3.24 | (2.56 - 4.1) | 2.5E-22 | 292 | 125 |
| Ibuprofen allergy | 3.15 | (2.19 - 4.52) | 5.3E-10 | 129 | 51 |
| Plantar warts (HPV) | 3.11 | (2.89 - 3.34) | 1.1E-207 | 3518 | 1387 |
| Macrolide allergy | 3.10 | (2.61 - 3.68) | 1.5E-38 | 572 | 230 |
| Azithromycin allergy | 3.07 | (2.31 - 4.07) | 6.9E-15 | 207 | 85 |
| Erythromycin allergy | 3.03 | (2.39 - 3.83) | 2.2E-20 | 304 | 119 |
| Cefaclor allergy | 3.00 | (2.37 - 3.81) | 1.4E-19 | 291 | 116 |
| Morphine hydromorphone allergy | 2.94 | (2.36 - 3.66) | 1.0E-21 | 346 | 135 |
| Hydrocodone allergy | 2.81 | (2.16 - 3.66) | 2.1E-14 | 239 | 93 |
| Codeine general allergy | 2.80 | (2.42 - 3.24) | 1.9E-43 | 808 | 307 |
| Opioid allergy | 2.79 | (2.48 - 3.15) | 6.3E-63 | 1206 | 454 |
| Sulfa allergy | 2.79 | (2.58 - 3.01) | 9.1E-150 | 3097 | 1143 |
| Codeine allergy | 2.78 | (2.32 - 3.34) | 1.9E-28 | 519 | 197 |
| Augmentin allergy | 2.71 | (2 - 3.68) | 1.3E-10 | 184 | 71 |
| Morphine allergy | 2.71 | (2.14 - 3.43) | 7.9E-17 | 311 | 116 |
| Clavulanate allergy | 2.70 | (2 - 3.64) | 8.6E-11 | 190 | 73 |
| Doxycycline allergy | 2.68 | (1.96 - 3.66) | 6.5E-10 | 175 | 67 |
| Acetaminophen allergy | 2.61 | (2 - 3.4) | 2.0E-12 | 239 | 88 |
| Penicillin allergy | 2.52 | (2.33 - 2.72) | 1.2E-122 | 3228 | 1109 |
| Vicodin allergy | 2.50 | (1.7 - 3.66) | 3.0E-06 | 116 | 42 |
| Cephalosporin specific allergy | 2.44 | (1.7 - 3.51) | 1.6E-06 | 133 | 47 |
| Chronic fatigue syndrome | 2.40 | (1.78 - 3.22) | 6.6E-09 | 202 | 70 |
| Amoxicillin allergy | 2.36 | (2.09 - 2.66) | 6.2E-45 | 1267 | 436 |
| Other inflammatory bowel disease (IBD) indeterminate colitis | 2.35 | (1.97 - 2.81) | 1.6E-21 | 582 | 196 |
| Sulfamethoxazole allergy | 2.29 | (1.88 - 2.78) | 8.9E-17 | 469 | 162 |
| Amoxicillin specific allergy | 2.29 | (2.01 - 2.6) | 2.6E-36 | 1096 | 369 |
| Trimethoprim with bactrim allergy | 2.27 | (1.87 - 2.77) | 2.7E-16 | 462 | 159 |
| Trimethoprim allergy | 2.24 | (1.83 - 2.74) | 3.7E-15 | 446 | 152 |
| Rosacea (acne rosacea) erythematotelangiectatic rosacea (ETR) | 2.19 | (1.98 - 2.43) | 1.9E-52 | 1865 | 606 |

| | | | | | |
|---|---|---|---|---|---|
| Bactrim allergy | 2.19 | (1.77 - 2.72) | 1.2E-12 | 386 | 130 |
| Hernias | 2.18 | (1.88 - 2.54) | 5.2E-24 | 824 | 258 |
| Ulcer peptic gastric or duodenal | 2.10 | (1.83 - 2.42) | 4.2E-25 | 946 | 298 |
| Claustrophobia | 2.04 | (1.8 - 2.31) | 1.3E-28 | 1212 | 376 |
| Skin condition | 2.03 | (1.93 - 2.13) | 1.0E-168 | 9651 | 2784 |
| Irritable bowel syndrome (IBS) spastic colon | 1.95 | (1.81 - 2.09) | 6.4E-75 | 4164 | 1219 |
| Bronchitis | 1.89 | (1.82 - 1.97) | 9.2E-218 | 27855 | 6635 |
| Fibromyalgia | 1.86 | (1.47 - 2.34) | 2.3E-07 | 347 | 102 |
| Digestive disorder | 1.85 | (1.75 - 1.95) | 2.1E-106 | 7867 | 2151 |
| Osteoarthritis (OA) degenerativejoint disease | 1.83 | (1.68 - 2) | 1.2E-41 | 2848 | 786 |
| Frequent heartburn | 1.77 | (1.69 - 1.86) | 7.1E-114 | 10600 | 2749 |
| Flu (respiratory) | 1.72 | (1.65 - 1.79) | 5.0E-143 | 41859 | 8867 |
| Migraines | 1.72 | (1.53 - 1.93) | 5.9E-20 | 1518 | 425 |
| Psoriasis | 1.68 | (1.51 - 1.87) | 2.0E-22 | 1920 | 517 |
| OCD (obsessive complusive disorder) | 1.68 | (1.53 - 1.83) | 3.5E-30 | 2757 | 738 |
| Raynaud's syndrome | 1.67 | (1.39 - 2) | 3.3E-08 | 604 | 165 |
| Anxiety | 1.65 | (1.58 - 1.72) | 1.6E-114 | 19891 | 4682 |
| Appendicitis | 1.64 | (1.37 - 1.96) | 5.5E-08 | 653 | 176 |
| Other cardiac arrhythmias (eg SVT) racing of the heart | 1.62 | (1.42 - 1.85) | 6.3E-13 | 1222 | 320 |
| ADHD/ADD | 1.59 | (1.5 - 1.69) | 2.5E-53 | 7113 | 1767 |
| Pneumonia | 1.58 | (1.51 - 1.65) | 6.1E-94 | 16127 | 3815 |
| Mental health/behavioral health condition | 1.57 | (1.5 - 1.63) | 3.8E-102 | 25186 | 5642 |
| Depression | 1.56 | (1.49 - 1.63) | 2.6E-92 | 19092 | 4402 |
| Childhood ear infection | 1.45 | (1.39 - 1.51) | 6.0E-73 | 36629 | 7547 |
| Yeast infection (candidiasis) | 1.44 | (1.38 - 1.5) | 1.9E-61 | 27312 | 5874 |
| Neurological disorder | 1.43 | (1.31 - 1.56) | 6.4E-15 | 2848 | 684 |
| Strep throat (streptococcal pharyngitis) | 1.41 | (1.35 - 1.48) | 1.0E-49 | 49317 | 9614 |
| Mononucleosis (EBV) | 1.39 | (1.33 - 1.46) | 1.3E-40 | 12418 | 2783 |
| Scarlet fever | 1.36 | (1.23 - 1.51) | 4.0E-09 | 2210 | 510 |
| Alcoholism | 1.36 | (1.19 - 1.55) | 5.3E-06 | 1338 | 293 |
| Hand foot and mouth disease (HFMD) Coxsackie | 1.36 | (1.26 - 1.47) | 3.1E-15 | 4189 | 965 |
| Scoliosis | 1.35 | (1.25 - 1.46) | 3.1E-14 | 4080 | 942 |
| Autoimmune disease | 1.34 | (1.27 - 1.42) | 2.2E-25 | 8733 | 1950 |
| Gallstones (cholelithiasis) | 1.33 | (1.24 - 1.44) | 1.5E-13 | 4305 | 965 |
| Heart condition | 1.32 | (1.21 - 1.44) | 4.4E-10 | 3246 | 716 |
| Chickenpox (varicella) | 1.28 | (1.22 - 1.34) | 1.0E-24 | 50607 | 9578 |
| Shingles | 1.27 | (1.19 - 1.35) | 1.3E-12 | 6262 | 1338 |
| Urinary Tract Infection (UTI) bladder infection | 1.26 | (1.2 - 1.31) | 1.3E-24 | 35934 | 7172 |

| | | | | | |
|---|---|---|---|---|---|
| Kidney stones (renal lithiasis) nephrolithiasis | 1.18 | (1.1 - 1.27) | 7.6E-06 | 5020 | 1006 |
| Cold sores | 1.17 | (1.12 - 1.22) | 1.9E-13 | 21453 | 4223 |
| *Contact dermatitis due to animal dander* | | | | 3591 | |
| Allergic rhinitis | 31.55 | (28.71 - 34.67) | < 2.2E-16 | 12779 | 3029 |
| Seasonal allergies to pollen | 19.39 | (17.11 - 21.97) | < 2.2E-16 | 28692 | 3311 |
| Contact dermatitis due to plants (except food) | 19.01 | (17.56 - 20.58) | < 2.2E-16 | 4028 | 1526 |
| Contact dermatitis due to soaps or detergents | 17.36 | (16.07 - 18.74) | < 2.2E-16 | 4836 | 1694 |
| Contact dermatitis due to food in contact with the skin | 14.84 | (12.71 - 17.31) | 9.5E-257 | 723 | 323 |
| Contact dermatitis due to other chemical products | 12.35 | (11.27 - 13.53) | < 2.2E-16 | 2627 | 928 |
| Contact dermatitis due to dyes | 12.23 | (10.32 - 14.49) | 1.4E-183 | 609 | 246 |
| Contact dermatitis due to cosmetics | 12.19 | (11.17 - 13.31) | < 2.2E-16 | 3007 | 1038 |
| Egg allergy | 10.73 | (7.12 - 16.15) | 6.3E-30 | 104 | 39 |
| Contact dermatitis due to adhesives | 10.20 | (9.37 - 11.09) | < 2.2E-16 | 3620 | 1081 |
| Contact dermatitis due to drugs in contact with skin | 9.47 | (8.02 - 11.19) | 3.6E-154 | 666 | 226 |
| Contact dermatitis due to metals | 9.03 | (8.25 - 9.89) | < 2.2E-16 | 2928 | 871 |
| Chemical sensitivities (eg perfumes) | 9.03 | (8.34 - 9.78) | < 2.2E-16 | 4763 | 1223 |
| Contact dermatitis due to latex | 7.99 | (7.09 - 9) | 4.3E-256 | 1499 | 438 |
| Asthma | 7.34 | (6.83 - 7.89) | < 2.2E-16 | 8268 | 1628 |
| Tree nuts and seeds allergy | 7.16 | (5.91 - 8.68) | 6.5E-90 | 560 | 162 |
| Peanut allergy | 7.09 | (5.54 - 9.07) | 7.3E-55 | 342 | 99 |
| Fish allergy | 6.80 | (4.87 - 9.5) | 2.8E-29 | 194 | 50 |
| Hives (urticaria) | 6.54 | (6 - 7.12) | < 2.2E-16 | 4014 | 916 |
| Food allergies or sensitivities | 6.33 | (5.9 - 6.8) | < 2.2E-16 | 9671 | 1690 |
| Eczema | 5.98 | (5.54 - 6.45) | < 2.2E-16 | 6466 | 1252 |
| Milk and dairy allergy | 5.90 | (4.09 - 8.5) | 2.0E-21 | 163 | 39 |
| Insect bites | 5.81 | (5.29 - 6.37) | 7.0E-303 | 3323 | 727 |
| Shellfish allergy | 5.47 | (4.55 - 6.57) | 9.6E-74 | 735 | 158 |
| Fruits and vegetables allergy | 5.02 | (4.08 - 6.18) | 2.8E-52 | 557 | 122 |
| Food additives allergy | 4.78 | (3.28 - 6.96) | 3.6E-16 | 177 | 38 |
| Lactose intolerance | 4.60 | (4.19 - 5.05) | 3.9E-228 | 3800 | 689 |
| Gluten intolerance (non-celiac) | 3.93 | (3.41 - 4.52) | 2.4E-81 | 1516 | 261 |
| Bee sting | 3.91 | (3.49 - 4.37) | 1.9E-124 | 2683 | 423 |
| Macrolide allergy | 3.69 | (2.94 - 4.62) | 1.5E-29 | 572 | 95 |
| Azithromycin allergy | 3.68 | (2.56 - 5.3) | 2.1E-12 | 207 | 37 |
| Nitrofurantoin allergy | 3.60 | (2.13 - 6.09) | 1.7E-06 | 102 | 17 |
| Erythromycin allergy | 3.56 | (2.59 - 4.89) | 5.5E-15 | 304 | 47 |
| Drug allergy | 3.55 | (3.3 - 3.82) | 6.4E-253 | 10734 | 1326 |
| Codeine allergy | 3.50 | (2.74 - 4.47) | 8.9E-24 | 519 | 81 |

| | | | | | |
|---|---|---|---|---|---|
| Plantar warts (HPV) | 3.47 | (3.13 - 3.85) | 1.2E-123 | 3518 | 518 |
| Morphine hydromorphone allergy | 3.47 | (2.58 - 4.65) | 1.4E-16 | 346 | 54 |
| NSAID (nonsteroidal anti-inflammatory drug) allergy | 3.45 | (2.57 - 4.65) | 2.9E-16 | 361 | 54 |
| Opioid allergy | 3.42 | (2.9 - 4.03) | 1.3E-48 | 1206 | 181 |
| Codeine general allergy | 3.38 | (2.77 - 4.13) | 3.3E-33 | 808 | 122 |
| Ciprofloxacin allergy | 3.37 | (2.24 - 5.06) | 4.8E-09 | 185 | 30 |
| Alopecia (alopecia areata) spot baldness | 3.34 | (2.4 - 4.64) | 6.7E-13 | 292 | 44 |
| Morphine allergy | 3.33 | (2.43 - 4.56) | 6.2E-14 | 311 | 47 |
| Fluoroquinolone allergy | 3.18 | (2.29 - 4.41) | 4.2E-12 | 306 | 45 |
| Aspirin allergy | 3.06 | (1.96 - 4.77) | 8.1E-07 | 176 | 24 |
| Clindamycin allergy | 3.05 | (1.89 - 4.91) | 4.9E-06 | 143 | 22 |
| Hydrocodone allergy | 3.00 | (2.08 - 4.33) | 4.1E-09 | 239 | 34 |
| Acetaminophen allergy | 2.86 | (1.96 - 4.16) | 4.5E-08 | 239 | 32 |
| Other inflammatory bowel disease (IBD) indeterminate colitis | 2.75 | (2.15 - 3.54) | 1.8E-15 | 582 | 75 |
| Penicillin allergy | 2.68 | | 6.3E-63 | 3228 | 381 |
| Claustrophobia | 2.66 | (2.23 - 3.16) | 4.8E-28 | 1212 | 156 |
| Sulfa allergy | 2.64 | (2.35 - 2.97) | 4.6E-60 | 3097 | 370 |
| Cefaclor allergy | 2.59 | (1.82 - 3.68) | 1.3E-07 | 291 | 36 |
| Penicillin class allergy | 2.58 | (2.33 - 2.86) | 2.0E-73 | 4315 | 494 |
| Ulcer peptic gastric or duodenal | 2.56 | (2.09 - 3.13) | 9.4E-20 | 946 | 114 |
| Rosacea (acne rosacea) erythematotelangiectatic rosacea (ETR) | 2.56 | (2.21 - 2.96) | 8.3E-36 | 1865 | 221 |
| Skin condition | 2.53 | (2.34 - 2.74) | 4.1E-121 | 9651 | 1007 |
| Cephalexin allergy | 2.47 | (1.68 - 3.63) | 4.0E-06 | 260 | 33 |
| Amoxicillin allergy | 2.27 | (1.9 - 2.72) | 2.7E-19 | 1267 | 145 |
| Hernias | 2.26 | (1.78 - 2.86) | 2.0E-11 | 824 | 78 |
| Amoxicillin specific allergy | 2.24 | (1.85 - 2.72) | 2.1E-16 | 1096 | 124 |
| Migraines | 2.17 | (1.84 - 2.57) | 1.3E-19 | 1518 | 166 |
| Bronchitis | 2.16 | (2.02 - 2.32) | 1.5E-103 | 27855 | 2055 |
| Sulfamethoxazole allergy | 2.11 | (1.56 - 2.84) | 1.1E-06 | 469 | 51 |
| Appendicitis | 2.10 | (1.61 - 2.73) | 3.6E-08 | 653 | 65 |
| Irritable bowel syndrome (IBS) spastic colon | 2.10 | (1.88 - 2.34) | 3.3E-40 | 4164 | 418 |
| Trimethoprim with bactrim allergy | 2.10 | (1.55 - 2.84) | 1.6E-06 | 462 | 50 |
| Trimethoprim allergy | 2.08 | (1.53 - 2.83) | 3.4E-06 | 446 | 48 |
| Frequent heartburn | 2.00 | (1.85 - 2.17) | 8.2E-64 | 10600 | 892 |
| Osteoarthritis (OA) degenerative joint disease | 1.99 | (1.72 - 2.29) | 5.0E-21 | 2848 | 242 |
| Flu (respiratory) | 1.96 | (1.81 - 2.11) | 1.5E-65 | 41859 | 2634 |
| OCD (obsessive compulsive disorder) | 1.95 | (1.71 - 2.23) | 7.5E-23 | 2757 | 273 |
| Digestive disorder | 1.93 | (1.77 - 2.11) | 3.5E-48 | 7867 | 690 |

| | | | | | |
|---|---|---|---|---|---|
| Raynaud's syndrome | 1.90 | (1.45 - 2.5) | 4.1E-06 | 604 | 59 |
| Anxiety | 1.88 | (1.75 - 2.02) | 2.0E-66 | 19891 | 1565 |
| ADHD/ADD | 1.84 | (1.68 - 2.02) | 9.3E-38 | 7113 | 630 |
| Pneumonia | 1.79 | (1.67 - 1.93) | 4.5E-55 | 16127 | 1214 |
| Depression | 1.78 | (1.66 - 1.92) | 1.1E-56 | 19092 | 1456 |
| Other cardiac arrhythmias (eg SVT) racing of the heart | 1.78 | (1.45 - 2.19) | 4.4E-08 | 1222 | 108 |
| Mental health/behavioral health condition | 1.77 | (1.65 - 1.9) | 6.9E-57 | 25186 | 1824 |
| Yeast infection (candidiasis) | 1.73 | (1.6 - 1.86) | 7.0E-48 | 27312 | 1937 |
| Neurological disorder | 1.71 | (1.49 - 1.96) | 5.2E-14 | 2848 | 240 |
| Alcoholism | 1.66 | (1.34 - 2.05) | 2.3E-06 | 1338 | 100 |
| Hand foot and mouth disease (HFMD) Coxsackie | 1.56 | (1.38 - 1.76) | 5.0E-13 | 4189 | 332 |
| Strep throat (streptococcal pharyngitis) | 1.54 | (1.42 - 1.68) | 3.0E-24 | 49317 | 2806 |
| Childhood ear infection | 1.50 | (1.4 - 1.62) | 4.7E-29 | 36629 | 2226 |
| Gallstones (cholelithiasis) | 1.50 | (1.32 - 1.69) | 1.9E-10 | 4305 | 313 |
| Psoriasis | 1.50 | (1.25 - 1.79) | 9.4E-06 | 1920 | 145 |
| Heart condition | 1.48 | (1.28 - 1.71) | 7.9E-08 | 3246 | 224 |
| Scarlet fever | 1.48 | (1.25 - 1.75) | 4.5E-06 | 2210 | 161 |
| Shingles | 1.41 | (1.26 - 1.57) | 8.5E-10 | 6262 | 406 |
| Mononucleosis (EBV) | 1.39 | (1.28 - 1.51) | 7.4E-15 | 12418 | 818 |
| Urinary Tract Infection (UTI) bladder infection | 1.37 | (1.27 - 1.48) | 4.5E-16 | 35934 | 2240 |
| Autoimmune disease | 1.36 | (1.23 - 1.49) | 2.5E-10 | 8733 | 576 |
| Scoliosis | 1.35 | (1.19 - 1.53) | 4.7E-06 | 4080 | 290 |
| Chickenpox (varicella) | 1.28 | (1.18 - 1.39) | 7.4E-09 | 50607 | 2655 |
| Cold sores | 1.26 | (1.17 - 1.35) | 3.5E-10 | 21453 | 1249 |
| *Hashimoto's disease* | | | | 1854 | |
| Raynaud's syndrome | 11.26 | (9.3 - 13.63) | 6.4E-136 | 604 | 160 |
| Chronic fatigue syndrome | 8.65 | (6.09 - 12.29) | 2.3E-33 | 202 | 44 |
| Graves' disease | 6.66 | (5 - 8.88) | 3.3E-38 | 345 | 62 |
| Fibromyalgia | 5.88 | (4.37 - 7.92) | 1.7E-31 | 347 | 54 |
| Sjögren's syndrome | 5.27 | (3.39 - 8.19) | 1.5E-13 | 163 | 25 |
| Lupus | 5.00 | (3.03 - 8.25) | 3.3E-10 | 139 | 19 |
| Celiac disease | 4.84 | (3.87 - 6.07) | 6.3E-43 | 786 | 92 |
| Vitiligo | 3.91 | (2.65 - 5.77) | 7.3E-12 | 325 | 31 |
| Rheumatoid arthritis | 3.78 | (2.68 - 5.33) | 3.1E-14 | 360 | 40 |
| Other cancer | 3.38 | (2.49 - 4.6) | 7.7E-15 | 535 | 49 |
| Gluten intolerance (non-celiac) | 2.71 | (2.21 - 3.32) | 7.7E-22 | 1516 | 109 |
| Psoriasis | 2.18 | (1.77 - 2.67) | 9.6E-14 | 1920 | 107 |
| Other cardiac arrhythmias (eg SVT) racing of the heart | 1.93 | (1.49 - 2.51) | 9.4E-07 | 1222 | 63 |

| | | | | | |
|---|---|---|---|---|---|
| Hives (urticaria) | 1.83 | (1.57 - 2.14) | 8.7E-15 | 4014 | 200 |
| Digestive disorder | 1.62 | (1.43 - 1.83) | 3.6E-14 | 7867 | 327 |
| Neurological disorder | 1.59 | (1.31 - 1.92) | 2.0E-06 | 2848 | 123 |
| Urinary Tract Infection (UTI) bladder infection | 1.48 | (1.33 - 1.64) | 2.8E-13 | 35934 | 1268 |
| Yeast infection (candidiasis) | 1.45 | (1.31 - 1.59) | 1.0E-13 | 27312 | 1029 |
| Bronchitis | 1.44 | (1.31 - 1.58) | 4.6E-14 | 27855 | 941 |
| Food allergies or sensitivities | 1.43 | (1.27 - 1.61) | 6.7E-09 | 9671 | 353 |
| Flu (respiratory) | 1.41 | (1.28 - 1.56) | 9.8E-12 | 41859 | 1246 |
| Skin condition | 1.40 | (1.24 - 1.58) | 5.0E-08 | 9651 | 344 |
| Strep throat (streptococcal pharyngitis) | 1.40 | (1.25 - 1.57) | 6.6E-09 | 49317 | 1443 |
| Mononucleosis (EBV) | 1.38 | (1.24 - 1.54) | 6.2E-09 | 12418 | 463 |
| Asthma | 1.37 | (1.2 - 1.56) | 2.8E-06 | 8268 | 283 |

OR: Odds ration; CI: Confidence interval; all associations under p-value $\leq 0.00001$
