## Supplementary Table 5 for "Self-reported health history from 70,724 individuals reveals novel HLA associations with allergy and other frequently underreported conditions"

| **Supplementary Table 5.** *HLA-DRB1*04:01* association *vs*. cefaclor, ceclor, and all cephalosporins | | | |
| --- | --- | --- | --- |
|  | **OR** | **CI_97.5%_** | **p-value** |
| Cefaclor (n = 30) | 3.87 | (1.84 - 7.95) | 2.39E-04 |
| Ceclor (n = 257) | 3.71 | (2.89 - 4.75) | 4.89E-25 |
| Cefuroxime (n = 31) | 1.49 | (0.59 - 3.28) | 3.60E-01* |
| Cephalexin (n = 245) | 1.34 | (0.97 - 1.81) | 6.40E-02* |
| Cefinidir (n = 41) | 1.04 | (0.42 - 2.21) | 9.20E-01* |
| Cefprozil (n = 46) | 0.48 | (0.14 - 1.19) | 1.60E-01* |
| Cephalosporin specific (n = 128)^†^ | 1.42 | (0.92 - 2.13) | 1.00E-01* |
| Cephalosporin class (n = 758)^‡^ | 1.98 | (1.68 - 2.32) | 1.70E-17 |
| OR: odds ratio; CI: confidence interval. *Non-significant. ^†^Means that the respondents answered “cephalosporin” as a specific term for drug allergy. ^‡^All different cephalosporins merged in the same class. All analysis in EUR population. | | | |
